# Digital phenotyping of aortic stenosis-related remodeling reveals complementary structural, electrical, and hemodynamic signatures

**DOI:** 10.64898/2026.07.22.26358600

**Authors:** Wenjing Luo, Ryan B. Choi, Doris Yang, Lovedeep S. Dhingra, Philip M. Croon, Rohan Khera, Evangelos K. Oikonomou

## Abstract

Aortic stenosis (AS) is a heterogeneous disease of aging characterized by valvular calcification and distinct structural, electrical, and hemodynamic remodeling that are incompletely captured by any single diagnostic measure. Here we show that three AI-derived digital biomarkers resolve AS-related remodeling into complementary structural (cine-CMR Digital AS Severity Index, DASSi), electrical (AI-ECG), and hemodynamic (phase-contrast CMR peak aortic velocity) axes. Among 68,714 UK Biobank participants, all three biomarkers were independently associated with prevalent AS and prospectively predicted aortic valve replacement. Genetic and transcriptomic analyses of the digital phenotypes revealed partially distinct, heritable architectures: peak aortic velocity aligned closely with clinical AS genetics, whereas DASSi and AI-ECG defined a shared myocardial-remodeling axis largely independent of clinical AS susceptibility. These findings support AS as a multidimensional remodeling syndrome and establish a novel digital phenotyping framework for dissecting complex cardiovascular disease into complementary, biologically informative axes.

## INTRODUCTION

Aortic stenosis (AS) is the most common valvular heart disease of aging and a leading cause of heart failure, disability and cardiovascular mortality worldwide. Its prevalence increases with age, affecting approximately 1-2% of adults older than 65 years and more than 10% of those over 75 years.^1^ Although AS is traditionally defined by progressive obstruction of left ventricular outflow, it is a biologically and clinically heterogeneous disease.^2^ Patients with similar transvalvular gradients may differ substantially in leaflet calcification, fibrosis, myocardial remodeling and electrical abnormalities.^3,4^ These variable manifestations reflect a spectrum of remodeling processes driven by interacting mechanical, inflammatory, metabolic, and genetic influences, which stands in contrast to the unidimensional definition of disease presence and severity based on hemodynamic thresholds.^5^

Disease progression likewise varies across physiological domains. *Hemodynamic* changes are marked by increasing velocities and gradients across the aortic valve as it becomes progressively stenotic.^6^ In parallel, *structural* remodeling of the valve and myocardium includes leaflet calcification and fibrosis, left ventricular hypertrophy, and the development of diastolic dysfunction in response to chronic pressure overload.^7,8^ *Electrical* remodeling also emerges as myocardial architecture, extracellular matrix composition, and loading conditions evolve.^9^ Together, these changes shape heterogeneous patterns of disease progression that are not quantified by conventional assessments that rely on single modalities. Recent advances in artificial intelligence (AI) applied to echocardiography, electrocardiography, computed tomography, and cardiac magnetic resonance imaging now enable scalable, automated characterization of flow, structural remodeling, and electrophysiological patterns, creating new opportunities to define AS phenotypes more comprehensively and track disease trajectories over time.^10^

Here, we hypothesized that an AI-enabled digital phenotyping strategy could characterize AS-related remodeling through complementary biomarkers capturing distinct disease dimensions. By moving beyond unidimensional severity thresholds, this approach may identify early transitions from subclinical remodeling to overt disease. We then used genetic and transcriptomic analyses to clarify what each biomarker represents by examining its relationship to clinically adjudicated AS, shared remodeling pathways, and inherited disease susceptibility. This framework may improve the interpretation of continuous digital phenotypes in AS and provide a generalizable approach to other cardiovascular diseases characterized by heterogeneous progression.

## RESULTS

### Data source and study population

We conducted a *post hoc* analysis of prospectively collected data from the UK Biobank, a large prospective population-based cohort with deep phenotyping, longitudinal follow-up, and standardized multiparametric cardiovascular phenotyping.^11^ This unique, integrated imaging framework enables simultaneous, operator-independent quantification of structural, electrophysiological, and flow-based phenotypes within the same visit, minimizing confounding by indication inherent to clinically referred cohorts. After exclusions for withdrawal of consent, prior aortic valve replacement (AVR), incomplete or missing biomarker or imaging data (see **Supplementary Fig. 1**), a total of 68,714 individuals were included in the study. At the time of the study visit, the median age was 66 [IQR: 60–72] years, and 35,682 (51.9%) were female. A total of 285 participants (0.4%) had an existing diagnosis of AS based on in-hospital diagnosis codes entered prior to the visit date. Further demographic and clinical characteristics of the study population are summarized in **Table 1**.

**Table 1.** Table of baseline characteristics.

| **Variable** | **Value** | **Statistics** |
| --- | --- | --- |
| **Total counts (n)** |  | 68,714 |
| **Age, median [Q1, Q3]** |  | 66 [60, 72] |
| **Sex, n (%)** | Female | 35,682 (51.9) |
|  | Male | 33,032 (48.1) |
| **Ethnic Background, n (%)** | White | 66,209 (96.4) |
|  | Asian or Asian British | 1,009 (1.5) |
|  | Black or Black British | 523 (0.8) |
|  | Other | 399 (0.6) |
|  | Mixed | 358 (0.5) |
|  | Prefer not to say | 163 (0.2) |
|  | Do not know | 53 (0.1) |
| **Diabetes, n (%)** |  | 4,085 (5.9) |
| **Hypertension, n (%)** |  | 20,977 (30.5) |
| **Hyperlipidemia n (%)** |  | 15,804 (23.0) |
| **Known Aortic Stenosis at baseline, n (%)** |  | 285 (0.4) |

Values represent counts (percentages), or median (25<sup>th</sup>-75<sup>th</sup> percentiles), as appropriate.

### Complementary digital biomarkers of AS-related remodeling

To operationalize our definition of AS as a multidimensional phenotype, we derived three complementary digital biomarkers capturing *structural*, *electrical*, and *flow-related remodeling* in AS using standardized, non-invasive data streams (**Fig. 1**).

**Fig. 1.**
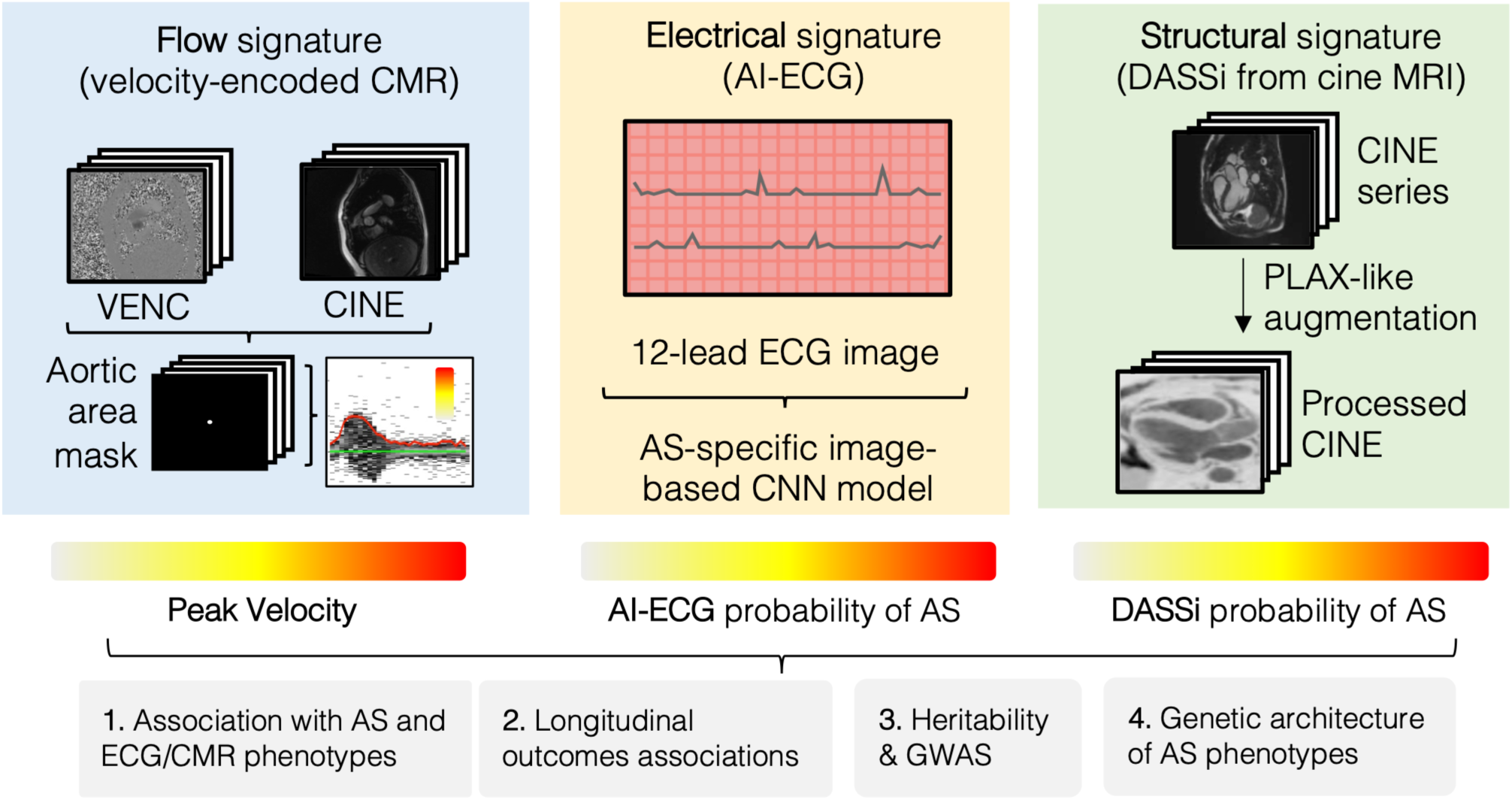
Multimodal digital profiling of aortic stenosis. Overview of a multimodal framework characterizing aortic stenosis (AS) through complementary electrophysiological, structural, and flow-based digital biomarkers. The electrophysiological signature is derived from a 12-lead ECG processed by an AI-enabled convolutional neural network (CNN), yielding a continuous probability of AS-related electrical remodeling. Structural remodeling is quantified using the Digital Aortic Stenosis Severity Index (DASSi), a deep learning model applied to PLAX-like long-axis cine cardiovascular magnetic resonance (CMR) images to capture valvular and myocardial remodeling independent of hemodynamics. Flow-related remodeling is assessed using an automated AI-based velocity-encoded CMR pipeline, in which the aortic lumen is segmented, and peak aortic velocity is extracted from the velocity-time profile. These complementary digital biomarkers are integrated to evaluate cross-sectional disease presence, longitudinal clinical outcomes, and downstream molecular associations, reframing AS as a multidimensional phenotype arising from partially independent remodeling processes. *Abbreviations:* AI, artificial intelligence; AS, aortic stenosis; CMR, cardiovascular magnetic resonance; CNN, convolutional neural network; DASSi, Digital Aortic Stenosis Severity Index; ECG, electrocardiogram; PLAX, parasternal long axis.

First, *structural remodeling* was quantified using the Digital AS Severity Index (DASSi), a deep learning-based phenotype originally trained on two-dimensional parasternal long-axis echocardiographic images without Doppler input to capture myocardial and valvular remodeling associated with moderate-to-severe AS.^4,12,13^ This approach has been validated across imaging platforms and was previously adapted to cine CMR by leveraging pre-processed three-chamber long-axis views,^4^ enabling generalizable structural phenotyping independent of hemodynamic measurements. DASSi has also been shown to be a surrogate of aortic valve calcium burden and calcification activity by computed tomography and ^18^F-NaF (sodium fluoride) positron emission tomography (PET) imaging,^14^ and is associated with parameters of diastolic function independent of left ventricular systolic function.^4^

Second, *electrophysiological remodeling* was assessed using an AI-enabled electrocardiogram (AI-ECG) score derived from the PRESENT-SHD framework,^15^ which was trained to discriminate the phenotype of moderate-to-severe AS directly from 12-lead ECGs. Like DASSi, we used the model’s output probability (ranging from 0 to 1) as a surrogate for the degree of AS-related electrophysiological remodeling learned by the model.

Finally, *flow-related hemodynamic remodeling* was quantified using peak aortic velocity, derived via an automated AI-based velocity-encoding pipeline applied to phase-contrast CMR.^16^ We adapted a previously validated U-Net segmentation model to delineate the aortic lumen at the sinotubular junction on magnitude images for each cardiac phase, with masks propagated to corresponding velocity-encoded frames (**Supplementary Fig. 2**).^16^ After excluding extreme pixel velocities (>3 standard deviations), the 99^th^ percentile velocity was computed per frame to generate a velocity-time curve, with the maximum value defined as peak aortic velocity.

Together, these biomarkers enabled us to capture distinct but complementary axes of AS-related remodeling, namely structural, electrical, and hemodynamic, enabling multimodal phenotyping within a unified protocolized framework.

### Digital AS-related remodeling biomarkers and cross-sectional AS presence

First, we assessed cross-sectional associations between the pre-defined, complementary digital biomarkers and clinically diagnosed AS in 68,714 UK Biobank participants. We confirmed that across modalities, digital biomarker values for all three axes of AS-related remodeling were higher among known AS cases versus individuals without known disease (**Fig. 2a-c**), and were associated with prevalent AS independently of each other, as well as age and sex (**Fig. 2d**). Although peak aortic velocity demonstrated the strongest association with prevalent AS (adj. OR of 2.92 [95% CI: 2.70–3.16] per each standard deviation [SD] increment), consistent with its central role in defining AS, both DASSi and AI-ECG maintained an independent association with cross-sectional disease (adj. OR of 1.45 [95% CI: 1.27–1.66] for DASSi and 1.41 [95% CI: 1.28–1.55] for AI-ECG) after explicitly accounting for peak aortic velocity, indicating that they capture complementary and nonredundant dimensions of AS-related remodeling beyond hemodynamic severity alone.

**Fig. 2.**
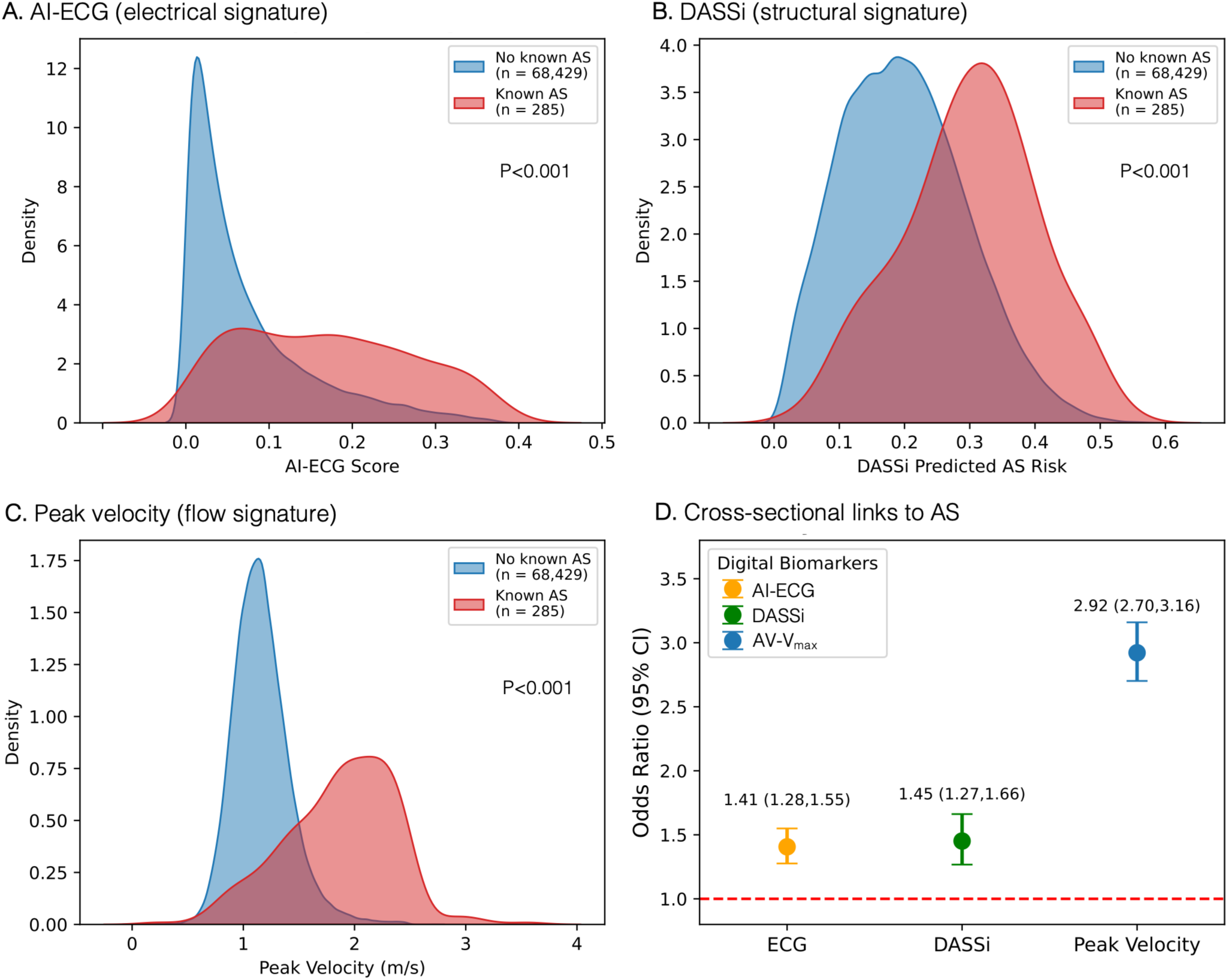
Cross-sectional associations between digital biomarkers and clinically diagnosed aortic stenosis. Distribution and associations of three complementary digital biomarkers with prevalent aortic stenosis (AS). (**A-B-C)**: Kernel density plots show the distribution of the electrophysiological (AI-ECG score) (**A**), structural (Digital AS Severity Index, DASSi) (**B**), and flow-based (peak aortic velocity) signatures (**C**), stratified by known AS status (AS, red; no known AS, blue). **(D)**: Adjusted odds ratios (adj. ORs) per standard deviation (SD) increments and 95% confidence intervals for prevalent AS per one standard deviation increase in each biomarker, estimated using multivariable logistic regression adjusted for age and sex. *Abbreviations:* adj., adjusted; AI-ECG, artificial intelligence-enabled electrocardiogram score; AS, aortic stenosis; CI, confidence interval; DASSi, Digital Aortic Stenosis Severity Index; OR, odds ratio; SD, standard deviation.

### Phenotypic selectivity of digital biomarkers across clinical, structural, and electrical domains

To further characterize the physiological domains captured by each biomarker beyond cross-sectional AS status, we evaluated their associations with broad clinical phenotypes and with quantitative CMR- and ECG-derived traits obtained at the same imaging visit.^17^ In phenome-wide association analyses (PheWAS), all three biomarkers showed enriched associations with cardiovascular risk factors, including obesity, diabetes mellitus, and hypercholesterolemia (**Supplementary Fig. 3**), consistent with shared relevance to cardiometabolic disease.^5^ By contrast, correlations with conventional imaging and ECG traits demonstrated clear domain selectivity (**Supplementary Fig. 4**). DASSi, a marker of structural remodeling, correlated more strongly with CMR-derived measures of cardiac structure, including left ventricular mass and atrial anatomy (e.g., LV mass, r = 0.36; left atrial maximum volume, r = 0.27), than did AI-ECG or peak aortic velocity. Conversely, AI-ECG showed stronger correlations with ECG-derived electrical features, including conduction and repolarization indices (e.g., PR interval, r = 0.32; QRS duration, r = 0.18; QTc, r = 0.13; all P < 0.001). Together, these findings suggest that although all three biomarkers relate to a common cardiovascular risk milieu, they capture distinct axes of AS-related remodeling across structural, electrophysiological, and hemodynamic domains.

### Digital AS-related remodeling biomarkers and AS progression

Next, we evaluated the prognostic associations of the three complementary digital biomarkers with incident AVR and mortality (**Fig. 3**). Over a median follow-up of 4.6 [IQR: 1.3–6.3] years, there were a total of 87 [0.1%] AVR events, and 1,161 [1.7%] deaths. In Cox proportional hazards models adjusted for age and sex, all three biomarkers independently predicted downstream AVR (**Fig. 3a**). Peak aortic velocity showed the strongest discrimination (adj. HR 2.68 [2.41–2.97] per SD; C-statistic of 0.94 [95% CI: 0.91– 0.96]), followed by DASSi and AI-ECG (adj. HR 1.90 [1.49–2.43] per SD; C-statistic 0.87 [95% CI: 0.84– 0.90] and adj. HR 1.60 [1.36–1.87] per SD; C-statistic 0.87 [95% CI: 0.83–0.90], respectively), whereas incorporating all 3 biomarkers boosted discrimination (C-statistic of 0.96 [95% CI: 0.94–0.98]) (**Fig. 3b**). Notably, incorporating all 3 biomarkers to a baseline model of age and sex did not substantially impact discrimination of mortality (C-statistic of 0.73) (**Fig. 3c-d**), consistent with the age-independent nature of these biomarkers.

**Fig. 3.**
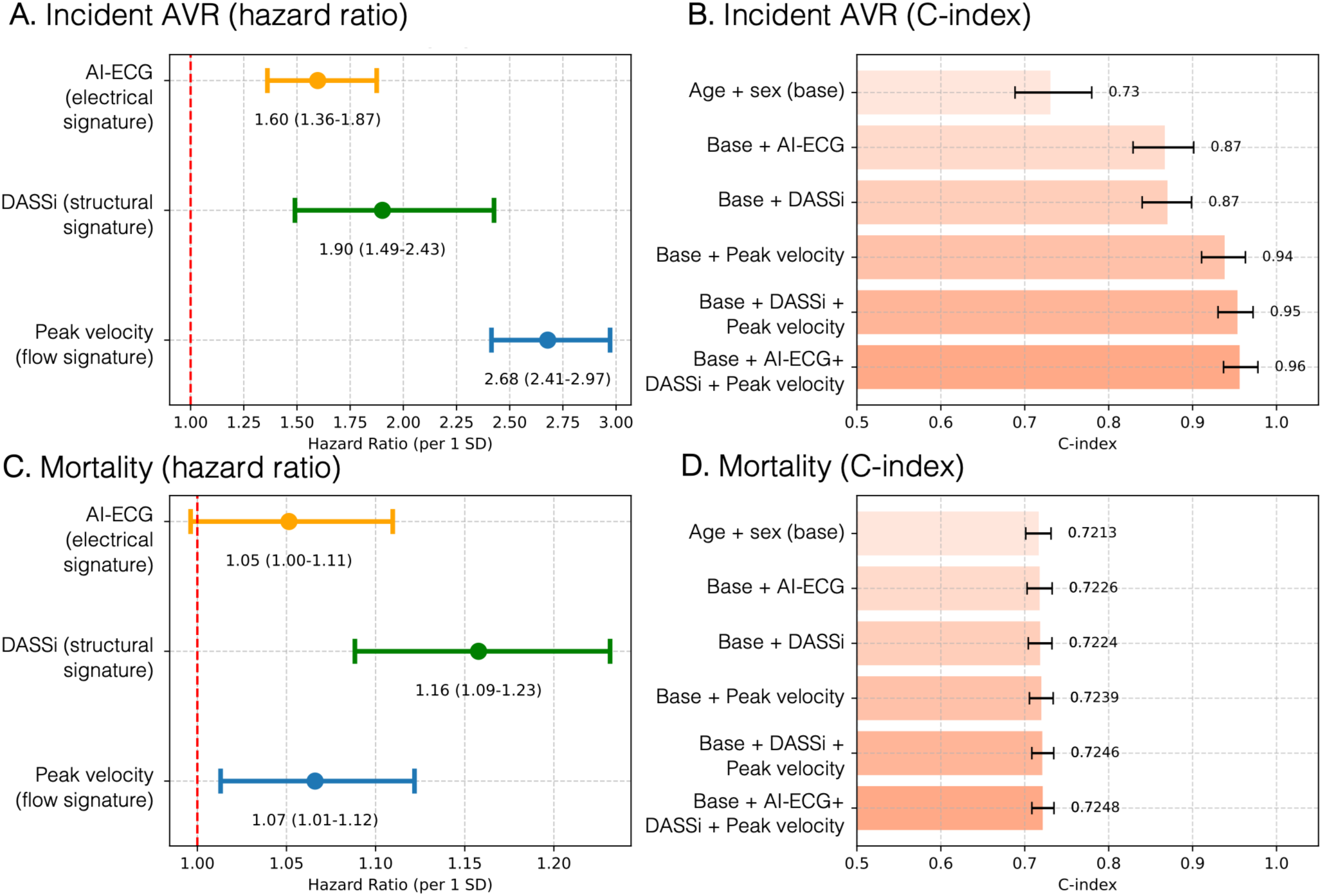
Prognostic associations of multimodal digital biomarkers with aortic valve replacement and mortality. (**A, C**) Hazard ratios (HRs) and 95% confidence intervals per 1-standard deviation (SD) increase in electrophysiological (AI-ECG), structural (Digital AS Severity Index, DASSi), and flow-based (peak aortic velocity) biomarkers for incident aortic valve replacement (AVR) (**A**) and all-cause mortality (**C**), estimated using Cox proportional hazards models adjusted for age and sex. All three biomarkers were independently associated with incident AVR, with peak aortic velocity showing the largest effect size, whereas associations with mortality were more modest. (**B, D**) Model discrimination for incident AVR (**B**) and all-cause mortality (**D**), assessed using Harrell’s concordance index (C-index) for age- and sex-adjusted models with sequential addition of individual or combined digital biomarkers. Error bars denote 95% confidence intervals. *Abbreviations:* adj., adjusted; AI-ECG, artificial intelligence-enabled electrocardiogram score; AS, aortic stenosis; AVR, aortic valve replacement; C-index, Harrell’s concordance index; CI, confidence interval; DASSi, Digital Aortic Stenosis Severity Index; HR, hazard ratio; SD, standard deviation.

### Subgroup analyses across hemodynamic strata

To further assess the value of multimodal phenotyping in clinically ambiguous scenarios, we stratified our study population based on a recently proposed peak aortic velocity threshold of 1.65 m/sec, a threshold that was selected to optimize the definition of mild AS across the population, as previously defined in the UK Biobank.^18^ Among individuals with low peak aortic velocities (<1.65 m/s, n=65,731 participants with 16 AVR events), DASSi remained a significant predictor of incident AVR, indicating prognostic signal despite sub-threshold flow measurements (**Supplementary Fig. 5a,b**). In contrast, among those with higher peak velocities (≥1.65 m/s, n=2,983 participants with 71 AVR events) suggestive of more advanced disease, both structural (DASSi) and electrophysiological (AI-ECG) biomarkers retained their independent association with incident AVR (**Supplementary Fig. 5c,d**). These findings support that reliance on flow-based thresholds incompletely captures disease trajectory and that multimodal digital biomarkers identify clinically relevant remodeling across the spectrum of AS severity.

### Genome-wide association of digital AS-related remodeling biomarkers

To characterize the genetic basis of these phenotypes, we performed genome-wide association studies (GWAS) for each continuous biomarker in 48,568 unrelated participants passing quality control (see **Methods**). Genomic inflation was well controlled (*λ*_GC_ = 1.016, 1.021, and 1.027 for DASSi, AI-ECG, and peak aortic velocity, respectively), with departure from the null confined to the extreme tail of the quantile-quantile plots (**Supplementary Fig. 6**), consistent with polygenic signal rather than confounding. Because three correlated phenotypes were analyzed, we report lead loci at both the conventional genome-wide threshold (P < 5 × 10^-8^) and a conservative phenotype-level Bonferroni threshold (P < 1.67 × 10^-8^): peak aortic velocity and AI-ECG each yielded 15 lead loci (13 each surviving the stricter threshold), whereas DASSi yielded no genome-wide-significant locus (**Fig. 4** and **Supplementary Table 1**). The 8p23.1 and 17q21.31 inversion regions are each reported as a single locus, represented by their most significant variant (rs7838131 and rs17608766, respectively) (***Methods***).

**Fig. 4.**
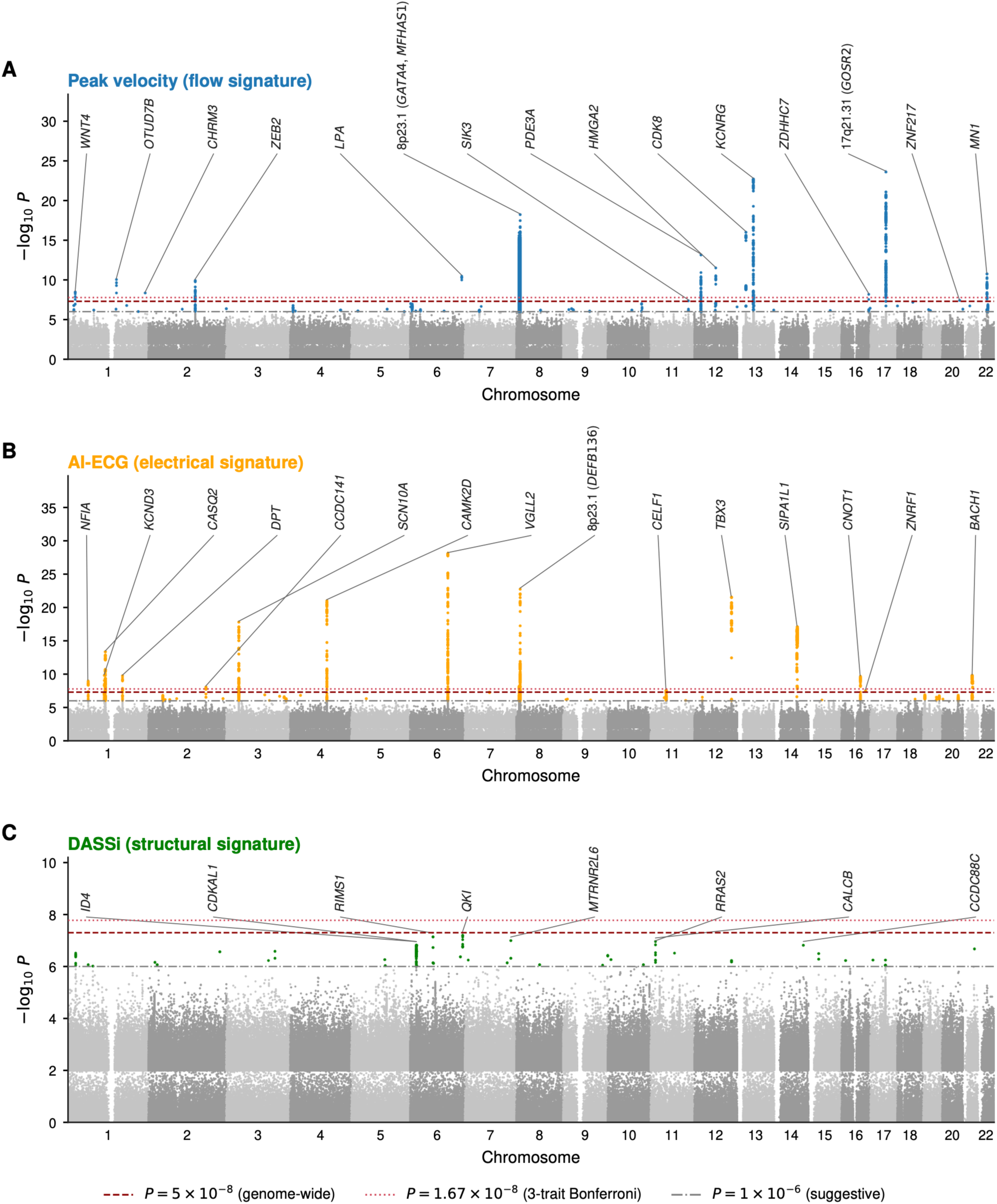
Genome-wide association landscape of the three aortic stenosis (AS)-related digital remodeling phenotypes. Manhattan plots are shown for each digital phenotype: (**A**) peak aortic velocity, **(B)** AI-ECG AS score, and (**C**) DASSi. Genome-wide-significant lead loci are labeled by nearest protein-coding gene; for DASSi, which reached no genome-wide-significant locus, the most significant suggestive loci are labelled. The two inversion regions are labelled by cytoband and reported as one locus each. Horizontal lines mark conventional genome-wide significance (P = 5 × 10⁻⁸), the conservative three-phenotype Bonferroni threshold (P = 1.67 × 10⁻⁸), and suggestive significance (P = 1 × 10⁻⁶). The corresponding quantile-quantile (QQ) plots and genomic-control inflation factors (λ_GC_) are shown in Supplementary Fig. 6. Abbreviations: AI-ECG, artificial intelligence-enabled electrocardiogram score; AS, aortic stenosis; DASSi, Digital Aortic Stenosis Severity Index; QQ, quantile-quantile; λ_GC_, genomic-control inflation factor.

The lead loci implicated distinct biology across phenotypes. Peak aortic velocity recovered established aortic valve and lipid genetics, including *LPA* (β = 0.077, P = 3.4 × 10^-11^),^19^ the 8p23.1 inversion locus (lead rs7838131, β = −0.057, P = 6.6 × 10^-19^),^20^ whose extended haplotype spans valve-development and previously implicated genes including *GATA4,*^20^ and *MFHAS1*,^21^ *CDK8* (β = 0.053, P = 1.0 × 10^-16^),^22^ *PDE3A* (β = −0.059, P = 7.4 × 10^-14^),^22,23^ *HMGA2* (β = −0.437, P = 3.4 × 10^-12^),^21,22^ and *KCNRG* (β = −0.063, P = 2.1 × 10^-23^).^24^ In contrast, the AI-ECG lead loci were expectedly dominated by a cardiac conduction and ion-channel program, including *SCN10A* (β = −0.053, P = 1.6 × 10^-18^), *TBX3* (β = 0.064, P = 2.8 × 10^-22^), *CAMK2D* (β = 0.064, P = 9.7 × 10^-22^), *CASQ2* (β = −0.045, P = 4.6 × 10^-14^) and *KCND3* (β = 0.046, P = 1.7 × 10^-10^), most not located at published AS loci and consistent with electrical remodeling related to but distinct from AS susceptibility. For DASSi, which yielded no genome-wide-significant locus, signals were only suggestive (P < 1 × 10^-6^). To place these loci against established AS genetics, we compared our lead variants with recently published AS studies (Small et al.^21^ and Kany et al.^22^) at the exact-variant, locus-window, and gene levels, and by direct look-up in their full summary statistics (**Supplementary Tables 2** and **3** and **Supplementary Fig. 7**). For instance, of the 15 peak aortic velocity lead loci, 11 lay within 500 kb of a published AS locus.^21,22^ On direct look-up in the published summary statistics, 12 were present in the UKB analysis by Kany et al.^22^ (all 12 with concordant effect direction; 8 at P < 5 × 10^-8^) and 14 in the calcific AS meta-analysis by Small et al.^21^

### Genetic architecture of digital AS-related remodeling biomarkers

We next examined the genetic architecture underlying these phenotypes. Correlations among the biomarkers were modest and concordant at the phenotypic and genetic levels (e.g., DASSi-vs-AI-ECG phenotypic r = 0.26 versus *r*_g_ = 0.27; **Fig. 5a,b**), while peak aortic velocity correlated minimally with either, resolving into a valve-hemodynamic axis and a shared myocardial-remodeling axis (**Fig. 5c**). Consistent with this separation, peak aortic velocity was strongly genetically correlated with published case-control GWAS of AS (*r*_g_ = 0.76 and 0.62), whereas AI-ECG and DASSi were at most weakly correlated with clinical AS (|r_g_| ≤ 0.16), markedly weaker than peak aortic velocity, indicating that only the hemodynamic biomarker is strongly genetically AS-proximal (**Fig. 5b** and **Supplementary Table 4**). SNP-based heritability was modest for all three phenotypes (peak aortic velocity h² = 0.18 [95% CI 0.10-0.26]; AI-ECG 0.15 [0.12-0.18]; DASSi 0.10 [0.08-0.12]; **Fig. 5d**). Together, these analyses position the biomarkers along a genetic spectrum, from a valve-proximal, AS-correlated axis (peak velocity) to a shared myocardial-remodeling axis (AI-ECG and DASSi) genetically independent of clinical AS.

**Fig. 5.**
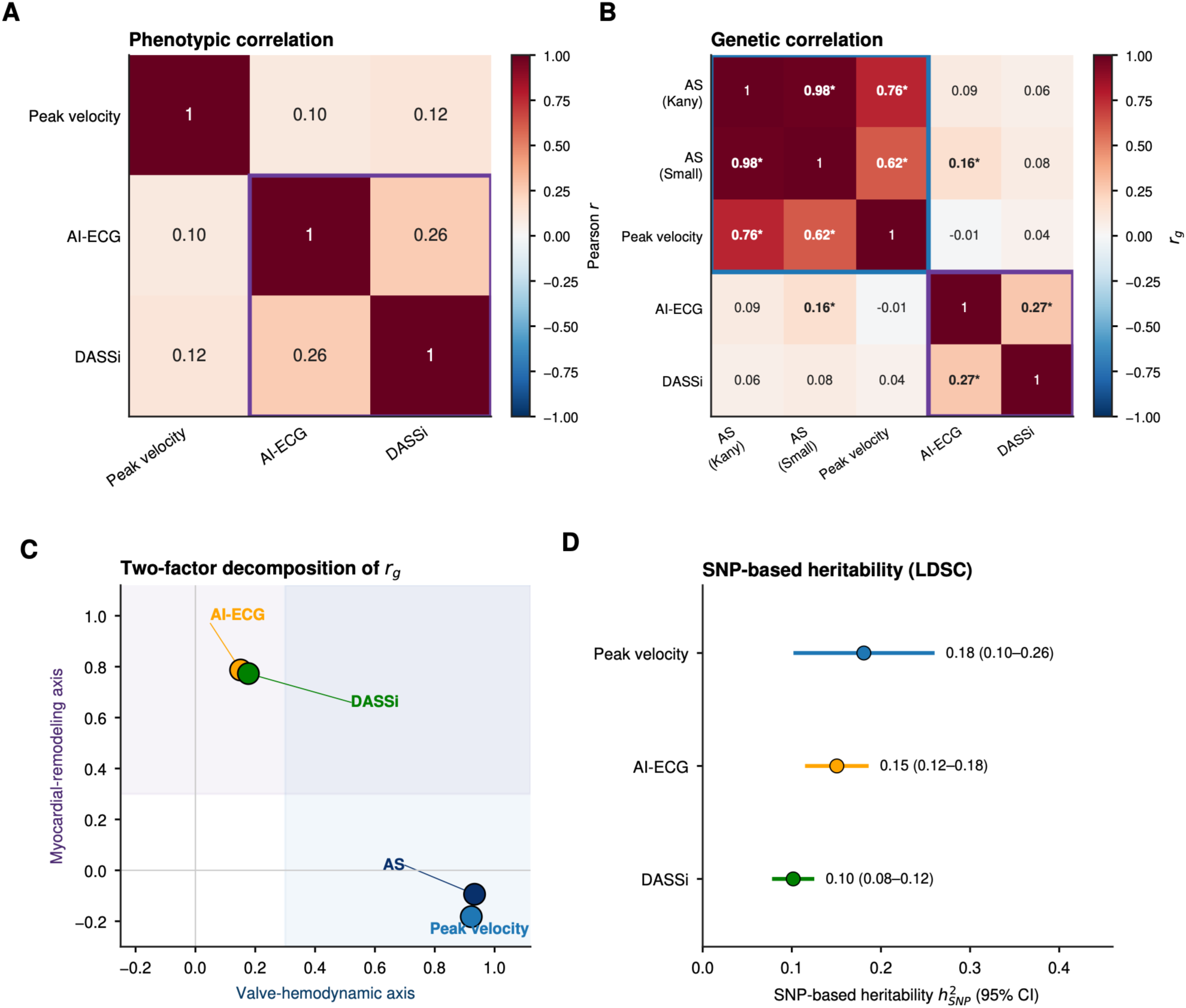
Genetic architecture of the digital remodeling phenotypes. (**A**) Phenotypic (Pearson) correlations among the three digital biomarkers. (**B**) Five-trait genetic-correlation matrix; asterisks mark pairs significant at P < 0.05. Boxes outline the valve-hemodynamic block (both AS references and peak velocity, blue) and the myocardial-remodeling block (AI-ECG and DASSi, purple). (**C**) Two-factor decomposition: Factor 1 (valve-hemodynamic) loads on the AS references and peak velocity; Factor 2 (myocardial-remodeling) loads on AI-ECG and DASSi. (**D**) SNP-based heritability with 95% confidence intervals. Notably, genetic correlations reflect shared genetic architecture, not causation. *Abbreviations:* AI-ECG, artificial intelligence-enabled electrocardiogram score; AS, aortic stenosis; CI, confidence interval; DASSi, Digital Aortic Stenosis Severity Index; LDSC, linkage-disequilibrium score regression; SNP, single-nucleotide polymorphism.

### Transcriptomic signatures of multimodal aortic stenosis phenotypes

Building on these observations, we performed transcriptome-wide association studies (TWAS) integrating GWAS summary statistics with tissue-specific genetically predicted gene expression models in relevant tissues (**Fig. 6**).^25^ The number of prioritized genes differed markedly across phenotypes (peak aortic velocity 135, AI-ECG 67, and DASSi 0 at a false-discovery rate [FDR] < 0.05; 11 for DASSi at FDR < 0.10) (**Supplementary Table 5**).

**Fig. 6.**
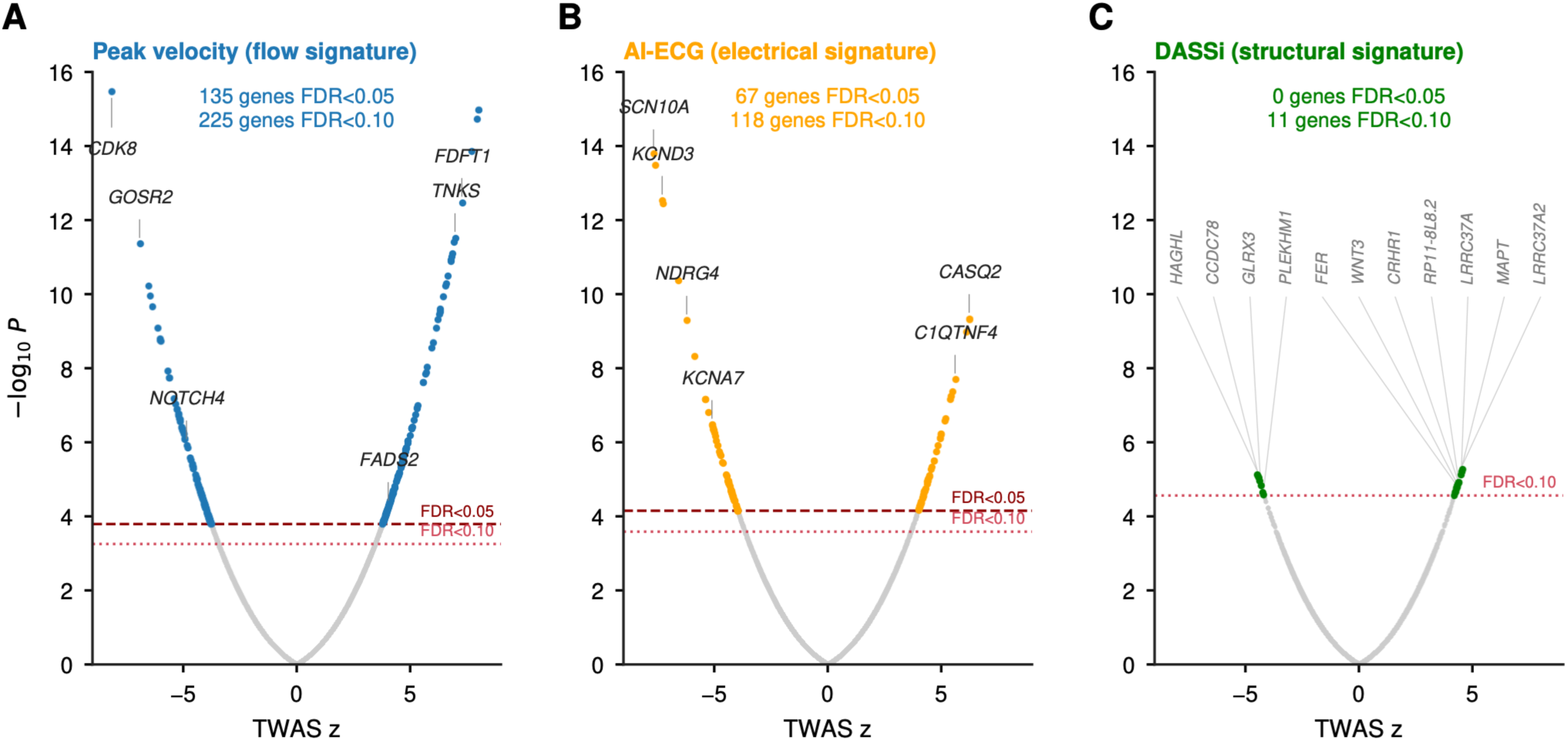
Transcriptome-wide association study (TWAS) across the three digital remodeling phenotypes. (**A**) Volcano plots of all gene-tissue associations for peak aortic velocity, (**B**) AI-ECG, and **(C)** DASSi, on a common −log10 P scale; the x-axis is the TWAS z (effect direction); grey points are non-significant; coloured points pass FDR < 0.05 (FDR < 0.10 for DASSi, which has no genes at FDR < 0.05); top genes are labelled. Peak velocity recovers established AS and aortic-valve genes (for example, *CDK8*, *FADS2*, and *NOTCH*4); AI-ECG is dominated by a cardiac conduction and ion-channel program (for example, *SCN10A*, *KCND3*, and *CASQ2*) largely distinct from AS; DASSi has no FDR < 0.05 genes and most of its suggestive genes lie in the 17q21.31 inversion region and are interpreted as provisional. *Abbreviations:* AI-ECG, artificial intelligence-enabled electrocardiogram score; AS, aortic stenosis; DASSi, Digital Aortic Stenosis Severity Index; FDR, false-discovery rate; TWAS, transcriptome-wide association study.

Peak aortic velocity identified established genes and pathways related to AS, aortic valve biology, and lipid metabolism, led by *CDK8* (z = −8.2, FDR = 3.2 × 10^-11^), with broad concordance with the independent human aortic valve TWAS by Small et al^21^ (21 shared genes, P = 2.9 × 10^-14^; P = 3.2 × 10⁻⁹ after collapsing each inversion region to a single unit; **Fig. 6a** and **Supplementary Table 6**). In contrast, the AI-ECG phenotype prioritized a cardiac conduction and ion channel program that did not localize to AS loci, such as *SCN10A* (z = −7.7, FDR = 1.5 × 10⁻⁹), as well as *KCND3*, *CASQ2*, and *NDRG4* (**Fig. 6b**). The AI-ECG signature showed limited overlap with the aortic valve TWAS, with three of the five shared genes lying within the 8p23.1 block (P = 0.11). Finally, the few DASSi-associated genes were borderline, and over half of them lay in the structurally complex 17q21.31 region (6 of 11 genes, all FDR ≈ 0.09), including *LRRC37A*, *LRRC37A2*, *MAPT* and *WNT3* (**Fig. 6c**),^26,27^ and this sparse signal did not exceed chance overlap with an independent human aortic valve TWAS^21^ (1 shared gene, *FER*; P = 0.11; **Supplementary Table 6**). We interpret these TWAS results as hypothesis-generating characterization of genetically regulated expression associated with the digital phenotypes rather than definitive effector-gene discovery.

### Colocalization across digital phenotypes and valve transcriptomic contextualization

Systematic colocalization across all prioritized GWAS-TWAS loci for the three phenotypes, reporting positive and negative results (PP.H4 and PP.H3; **Supplementary Table 7** and **Supplementary Figs. 8-10**), showed tissue-specific colocalization at coherent genes for peak aortic velocity and AI-ECG (for example, *CDK8* in cardiovascular tissues; *CASQ2* and *SCN10A* in heart). Of the six genes colocalizing for DASSi, three lie in the 17q21.31 inversion region, and three outside it (namely *CCDC78, HAGHL,* and *GLRX3*). Within 17q21.31, *WNT3* colocalized preferentially in cardiovascular tissues (PP.H4 ≈ 0.81-0.90), a more plausible signal within this complex locus, which is supported by prior evidence related to its role in osteoblast differentiation signaling in calcific AS.^26^

Finally, as supportive transcriptomic contextualization, we examined the prioritized peak-velocity gene set in independent public valve datasets. In a single-cell atlas of human aortic valves (82,356 cells, five donors, non-diseased to fibrocalcific; GSE220774),^28^ the set was expressed predominantly in valvular interstitial and endothelial populations rather than *immune* cells (88/118 genes peaking in structural cells; P = 6 × 10^-4^), and its expression tracked disease state in both populations. In independent bulk RNA-sequencing, the set was enriched among genes differentially expressed in calcific versus non-calcified valves (GSE199718;^29^ Fisher P = 2 × 10^-4^), with a concordant but non-significant trend in an AS-specific contrast of stenotic versus normal leaflets (GSE153555;^30^ Fisher *P* = 0.07) (**Supplementary Table 8).**

## DISCUSSION

In this study, we present a multimodal framework that characterizes AS as a heterogeneous disease, defined by complementary yet largely independent remodeling across structural, electrophysiological, and hemodynamic domains. Leveraging large-scale, protocolized data from the UK Biobank, we demonstrate that three digital biomarkers, a cine-CMR-derived structural index (DASSi),^4,12,13^ an AI-enabled electrocardiogram (AI-ECG) score,^15^ and flow-based peak aortic velocity,^16,18^ capture distinct yet complementary aspects of the AS spectrum, from cross-sectional phenotypes, to longitudinal progression and biology. Each biomarker was independently associated not only with cross-sectional disease presence, but also longitudinal clinical outcomes, while exhibiting domain-specific phenotypic, prognostic, and genetic signatures. Together, our findings support an actionable and translatable paradigm in which AS is not adequately described by a single severity axis but instead reflects the convergence of multiple remodeling processes that may be quantified digitally at scale.

Our findings extend and integrate prior work in several important ways. Traditional approaches to AS have largely focused on flow-derived metrics, particularly transvalvular velocity and gradient, as the dominant determinants of diagnosis and clinical decision-making.^31,32^ While these measures remain central, emerging evidence has highlighted substantial heterogeneity in myocardial remodeling, electrical conduction, and clinical outcomes among individuals with similar hemodynamic severity.^6^ Recent studies, including prior work introducing DASSi,^4,12,14^ demonstrated that structural remodeling quantified from imaging data provides prognostic information beyond conventional flow measures. Similarly, AI-based ECG models have been shown to detect subclinical structural heart disease from surface electrical signals.^15^ By integrating these approaches within a single population-based framework, our study demonstrates that these modalities capture complementary dimensions of AS. Importantly, the divergence observed at the phenotypic level is mirrored by partially distinct genetic and transcriptomic signatures, reinforcing the biological plausibility of a multimodal disease architecture.

This framework offers several conceptual and translational advantages and is aligned with the multimodal reasoning of clinical practice. First, it enables AS-related remodeling to be characterized along continuous digital axes rather than only as a binary diagnosis, increasing sensitivity to early or subclinical disease states, as has previously been shown for traditional hemodynamic metrics.^22^ Second, agreement or discordance across digital biomarkers may improve diagnostic precision by identifying individuals in whom multiple remodeling processes are already engaged, even when conventional thresholds are not met. Third, multimodal phenotyping may boost prognostic stratification, as demonstrated by incremental gains in discrimination for AVR when combining structural, electrophysiological, and flow signatures. Most importantly, this approach provides evidence to enhance disease phenotyping and candidate gene prioritization by enabling the dissection of disease heterogeneity into component processes that may be linked to distinct molecular pathways.

Beyond AS, this paradigm is broadly applicable to other forms of cardiomyopathy and complex cardiovascular diseases characterized by parallel remodeling across multiple domains. Conditions such as hypertrophic cardiomyopathy, heart failure with preserved ejection fraction, and infiltrative cardiomyopathies exhibit substantial heterogeneity in structural, electrical, and functional manifestations.^33,34^ Thus, multimodal digital phenotyping offers a scalable approach to decompose these conditions into biologically meaningful subphenotypes, with implications for diagnosis, prognosis, and therapeutic targeting. By moving beyond single-modality or threshold-based definitions, this framework aligns with emerging precision medicine efforts that seek to characterize disease as a dynamic process rather than a static label.

Certain limitations merit consideration. *First*, the UK Biobank is not fully representative of the general population and has otherwise limited ethnic diversity. Nevertheless, both DASSi and AI-ECG were previously trained and validated across racially and ethnically diverse populations,^4,12,15^ and were deployed in this cohort without further finetuning. *Second*, the genome-wide, transcriptome-wide, and colocalization analyses were performed on continuous, model-derived digital biomarkers rather than on clinically adjudicated AS or direct valve-tissue measurements and remain associative. Their genetic architecture may reflect correlated heritable traits, including myocardial size, conduction, cardiometabolic risk, and age-related remodeling, in addition to AS-related biology; consistent with this, only peak aortic velocity was strongly genetically correlated with clinically adjudicated AS. Recent large multi-ancestry AS GWAS, which implicate lipid metabolism, inflammation, cellular senescence, and adiposity in calcific AS, provide the appropriate reference for AS susceptibility loci,^21^ and we interpret our analyses as characterizing AS-related digital remodeling phenotypes rather than as definitive evidence of AS-specific causal mechanisms. Relatedly, two of the regions we report, namely 8p23.1 and 17q21.31, lie within common inversion polymorphisms, so their associations cannot be resolved into independent variants or assigned to individual effector genes from summary statistics alone. *Third*, the three phenotypes differ in their proximity to AS. Peak aortic velocity reproduces much of the established genetic architecture of AS; DASSi may partly reflect left ventricular hypertrophy or myocardial remodeling, an epiphenomenon more removed from the valve lesion itself and a convergent endpoint of diverse maladaptive responses. The value of the multimodal approach therefore lies not in any single biomarker but in resolving these distinct, complementary layers of AS-related remodeling. However, perturbational experiments and additional valve-tissue datasets are required to establish causal roles for any prioritized genes. *Fourth*, CMR-derived peak aortic velocity is not interchangeable with Doppler echocardiographic V_max_ and may underestimate eccentric or spatially heterogeneous jets. On the same note, the 1.65 m/s cut-point is a population-derived CMR threshold rather than a Doppler echocardiographic definition of mild AS,^18^ and the corresponding subgroup findings should therefore be interpreted as exploratory. *Fifth*, incident AVR is a treatment-dependent endpoint influenced by symptoms, comorbidity, competing mortality, referral patterns, and clinical decision-making, and the small number of AVR events, particularly in the lower velocity subgroup, warrants cautious interpretation of the subgroup discrimination estimates.

In conclusion, we show that AS-related remodeling can be characterized through the integration of complementary digital biomarkers capturing distinct electrical, structural, and flow-related remodeling processes. This digital biomarker framework moves beyond single-axis disease definitions, capturing the complex clinical entity of AS and reframing it as a heterogeneous, biologically layered condition that evolves along partially independent pathophysiological trajectories. By anchoring disease characterization to continuous, predictive digital phenotypes, this approach provides a scalable and generalizable paradigm for studying complex cardiovascular diseases and for linking clinical risk stratification to underlying biological mechanisms through digitally defined remodeling signatures.

## METHODS

### Study Population and Data Sources

The UK Biobank is a prospective population-based cohort comprising over 500,000 participants aged 40-69 years at enrollment. All analyses were conducted under the project ID #71033. For this post hoc analysis, we included participants who underwent CMR imaging as part of the UK Biobank imaging sub-study between 2014 and 2024.^11^ Among these, 68,714 individuals had complete and valid data, spanning velocity-encoded CMR videos, cine-CMR long-axis (3-chamber) views and 12-lead ECGs and were included in the primary analyses. Participants were excluded if they had withdrawn consent, had a history of AVR prior to imaging, had incomplete or poor-quality CMR data, or failed one or more automated image-processing pipelines. Race was self-reported and categorized as Asian (including Asian British or Chinese), Black (or Black British), White, or Other (including multiracial, unknown, or not reported).

### ECG and CMR imaging

Resting 12-lead ECGs were acquired as part of the protocolized UK Biobank baseline assessment using standardized acquisition protocols and digitally stored in raw waveform format. ECGs were sampled at a frequency of 500 Hz with a duration of 10 seconds per lead and were processed using automated quality-control procedures prior to being plotted and analyzed, as previously described.^35^ All CMR scans were acquired using a standardized 20-minute protocol on a Siemens 1.5-Tesla MAGNETOM Aera scanner (Siemens Healthineers, Erlangen, Germany).^36^ Phase-contrast flow imaging was performed perpendicular to the ascending aorta above the sinotubular junction to quantify aortic valve flow, and included paired magnitude reference images and velocity-encoded images generated using a planned velocity encoding (VENC) parameter, typically 2.5 m/s as recorded in the DICOM/CSA headers. Across the cardiac cycle, 30 retrospectively gated images were obtained with a slice thickness of 6 mm, an in-plane voxel size of 1.77 × 1.77 mm, and individualized temporal resolution based on heart rate. Where applicable, standard CMR measurements beyond DASSi were defined using the measurements provided by *Bai et al*.^17^

### Digital AS Severity index (DASSi)

DASSi was derived using a previously published deep learning model originally trained on two-dimensional parasternal long-axis (PLAX) echocardiographic images without Doppler input.^4,12^ To enable application within the UK Biobank CMR dataset, an automated pipeline retrieved raw cine CMR data and identified left ventricular outflow tract (LVOT) long-axis sequences corresponding to the three-chamber view. Cine frames were rotated, cropped to the cardiac silhouette, and converted to grayscale to approximate the appearance of PLAX echocardiographic images, as previously described.^4^ The pretrained model was then applied to each cine video to generate a continuous probability score ranging from 0 (lowest probability of severe AS) to 1 (highest probability), which was used as the DASSi structural biomarker.

### AI-ECG AS score

The AI-ECG score was generated using a previously validated deep learning model trained to detect moderate-to-severe AS from resting 12-lead ECG images.^15^ The model architecture and training procedures have been described in detail elsewhere and consist of a convolutional neural network (CNN) trained on ECG waveform images to learn AS-related electrophysiological features independent of imaging-derived severity metrics. Similar to DASSi, the model was applied in inference-only mode without retraining or fine-tuning on UK Biobank data. Raw ECG waveforms were converted to standardized image representations,^35^ and processed by the pretrained network to generate a continuous probability score reflecting the likelihood of AS-related electrical remodeling. This probability score was used as the AI-ECG digital biomarker in all analyses.

### Peak aortic velocity

To estimate the peak aortic velocity, we adapted an automated AI-based velocity-encoding pipeline applied to phase-contrast CMR data.^16^ More specifically, a previously validated U-Net segmentation model (DeepFlow) was applied to magnitude images to delineate the aortic cross-section at the sinotubular junction for each cardiac phase. Segmentation masks were then overlaid onto the corresponding velocity-encoded images to extract pixel-level velocity values (cm/s). The sign of the velocity encoding was determined based on the assumption that the total blood flow within one cardiac cycle is forward. To reduce noise, extreme velocity values exceeding three standard deviations from the frame-specific mean were excluded. For each cardiac phase, the 99^th^ percentile of remaining velocities was computed to generate a velocity-time curve across the cardiac cycle. The maximum value of this curve was defined as the participant’s peak aortic velocity and used as the flow-based digital biomarker.

### Epidemiological and Clinical Outcome Definitions

Prevalent AS and cardiometabolic comorbidities were identified using International Classification of Diseases (ICD), Tenth Revision (ICD10) hospital diagnosis codes recorded prior to or at the CMR exam (I35.0, I35.2). Aortic stenosis based on documented ICD codes was used to ensure accuracy. Prior AVR procedures were identified using Office of Population Censuses and Surveys Classification of Surgical Operations and Procedures, Fourth Revision (OPCS-4) procedural codes, as previously described.^4^ Participants with evidence of AVR before the CMR scan used for analysis were excluded. The primary outcome was time to incident AVR, and the secondary outcome was time to all-cause mortality. Time-to-event for AVR was defined as the interval from the CMR imaging date to the first recorded AVR procedure, whereas time to death was defined as the interval from the CMR date to the date of death from any cause, ascertained through linkage with national death registries. Participants without events were censored at the end of available follow-up.

### Phenome-Wide Association Studies

PheWAS studies were performed as previously described,^4,37^ using available packages,^38,39^ with ICD-9/10-derived diagnosis codes recorded before or on the day of the CMR/ECG visit collapsed into phenotypic codes (phecodes) and effect estimates derived from age and sex-adjusted linear regression models with each one of the three digital biomarkers treated as the dependent variable in three separate models.

### Genome-Wide Association Studies

Genome-wide association analyses were performed for continuous digital AS phenotypes using the UK Biobank genotype data. We restricted analyses to unrelated participants of White British ancestry to minimize population stratification, as defined by UK Biobank genetic ancestry inference. GWAS analyses were conducted using REGENIE (v2.1.1) under a two-step whole-genome regression framework optimized for large-scale biobank data. In step 1, a null model was fit using directly genotyped variants to account for polygenic background and relatedness. In step 2, imputed variants were tested using a linear regression framework assuming an additive genetic model. All models were adjusted for age at imaging, sex, and the first ten principal components of genetic ancestry. For genotype array data, variants on sex chromosomes were excluded, as were variants with minor allele frequency (MAF) < 0.01, minor allele count < 100, variant missingness > 0.1, or Hardy-Weinberg equilibrium *P* < 1 × 10⁻¹⁵. For imputed variants derived from the GEL imputation panel, variants on sex chromosomes were excluded, along with those with MAF < 1 × 10⁻⁴, minor allele count < 10, variant missingness > 0.1, or Hardy-Weinberg equilibrium *P* < 1 × 10⁻¹⁵. Genome-wide significance was defined as *P* < 5 × 10⁻⁸, with suggestive significance defined as *P* < 1 × 10⁻⁶. Because three correlated digital phenotypes were analysed, we additionally applied a conservative phenotype-level Bonferroni threshold of P < 1.67 × 10^-8^ (= 5 × 10^-8^ / 3) and indicate loci meeting this threshold. For each phenotype we generated Manhattan and quantile-quantile (QQ) plots and computed the genomic-control inflation factor (*λ*_GC_) to assess residual confounding. Approximately independent lead loci were defined by distance-based clumping, iteratively selecting the most significant remaining variant and removing all variants within ±500 kb on the same chromosome; this is an approximate, distance-based locus definition rather than linkage-disequilibrium-based fine-mapping. Lead loci were annotated to the nearest protein-coding gene. Two structurally complex regions, 8p23.1 and 17q21.31, lie within common inversion polymorphisms in which linkage disequilibrium extends across several megabases; each was therefore reported as a single locus per phenotype, represented by its most significant variant, with the remaining leads within them flagged in **Supplementary Table 1** and not counted separately. The summary statistics were used for transcriptome-wide association studies, as described below.

### Heritability, genetic and phenotypic correlations

We estimated SNP-based heritability (h²) and pairwise genetic correlations (*r*_g_) using LD score regression (ldsc v1.0.1), restricting REGENIE summary statistics to HapMap3 variants and using precomputed LD scores from 1000 Genomes Phase 3 European samples for both the reference panel and regression weights; 95% confidence intervals were derived from the LDSC standard errors. Genetic correlations were estimated among the three digital phenotypes and between each phenotype and two published meta-analyses of clinically adjudicated case-control AS (Kany et al.^22^ and Small et al.^21^). We additionally report the pairwise phenotypic (Pearson) correlations between the biomarkers. To characterize the shared structure, we performed a two-factor eigendecomposition of the five-trait genetic-correlation matrix and computed partial genetic correlations between phenotypes conditional on the others.

### Transcriptome-wide Association Studies

To prioritize candidate genes underlying digital AS phenotypes, we performed transcriptome-wide association studies (TWAS) using S-PrediXcan,^25^ integrating GWAS summary statistics with tissue-specific gene expression prediction models. GWAS summary statistics for each continuous digital biomarker (Digital AS Severity Index [DASSi], AI-ECG score, and peak aortic velocity) were generated using REGENIE and harmonized to the GRCh38 reference genome. Gene expression prediction models were obtained from GTEx v10 (https://gtexportal.org/home/)^40^ using mashr-based multivariate adaptive shrinkage,^41^ which leverages cross-tissue correlation to improve effect size estimation. Analyses focused on cardiovascular and metabolically relevant tissues, including left ventricle, atrial appendage, aorta, coronary artery, visceral adipose tissue, liver, and whole blood. GWAS summary statistics were aligned to MetaXcan-compatible variant identifiers (chr:position:ref:alt), with allele harmonization performed prior to analysis. S-PrediXcan was run separately for each digital biomarker across tissues, yielding gene-level association statistics. Multiple testing was addressed using Benjamini-Hochberg false discovery rate (FDR) correction within each GWAS; genes were considered significant at FDR < 0.05 and, more permissively, at FDR < 0.10 (both are reported), and prioritized genes were carried forward for downstream analyses. Gene-level association statistics were visualized using volcano plots displaying TWAS Z-scores and corresponding P values for each digital phenotype.

### Colocalization analyses

To distinguish shared causal signals from coincidental associations driven by linkage disequilibrium, we performed colocalization systematically across all prioritized GWAS-TWAS loci for all three digital phenotypes, rather than for a selected subset. For each gene locus, regional GWAS summary statistics (±200 kb around the lead variant) were aligned with GTEx cis-eQTL summary statistics for seven cardiovascular and metabolically relevant tissues and evaluated with the Bayesian coloc.abf framework. Loci were prioritized using phenotype-appropriate TWAS thresholds (peak velocity FDR < 0.05; AI-ECG and DASSi FDR < 0.10). We report the posterior probability of a single shared causal variant (PP.H4) and of distinct causal variants (PP.H3), and present all tested loci, including those without evidence of colocalization. Colocalization results are to be interpreted conservatively and used to support gene prioritization rather than definitive causal inference.

### Comparison with published AS loci and enrichment analyses

We compared our lead loci with two large published AS genetic studies (Small et al. multi-ancestry calcific-AS meta-analysis,^21^ and Kany et al.^22^) at three levels: exact variant (shared rsID or chr:position:ref:alt), locus window (a lead variant within ±500 kb of a published lead), and gene level (shared nearest or mapped gene). We additionally looked up each genome-wide-significant lead variant directly in the published full summary statistics, aligning effect sizes to our effect allele and recording effect-direction concordance. Enrichment of our loci among published AS loci was assessed with a binomial test against the genome-wide background expectation (the fraction of the genome within ±500 kb of any published AS lead). Cross-study concordance of prioritized genes was tested by comparing our TWAS-prioritized genes with the human aortic-valve TWAS reported by Small et al. using a hypergeometric test against the background of all TWAS-tested genes.^21^ Because genes within an inversion polymorphism are not independent units, the hypergeometric test was repeated as a sensitivity analysis in which all genes inside 8p23.1 or 17q21.31 were collapsed to a single unit per region, in the query set, the reference set and the tested-gene background.

### Valve transcriptomic contextualization

To place the prioritized genes in a tissue context, we examined their expression in human aortic valve transcriptomic datasets. Single-cell RNA-sequencing of human aortic valves spanning a disease-severity gradient (non-diseased to fibrocalcific) was obtained from GSE220774;^28^ cells were assigned to valvular interstitial, valvular endothelial, transitional, myeloid, and lymphoid populations, and canonical markers were used to confirm assignments (i.e., *DCN* and *ACTA2* for valvular interstitial cells, *PECAM1* and *VWF* for valvular endothelial cells, and *CD68* for myeloid cells). Counts were library-size normalized and aggregated to per-cell-type, per-donor pseudobulk. We tested whether the TWAS- and colocalization-prioritized genes (predominantly the AS-specific peak velocity gene set) were preferentially expressed in valve structural cells relative to immune cells (Mann-Whitney test against a matched background gene set) and whether their pseudobulk expression tracked the donor disease-severity gradient (Spearman correlation). We additionally tested their differential expression in independent bulk RNA-sequencing cohorts of calcific versus non-calcific aortic valves (GSE199718 and GSE153555) using rank-based enrichment.^29,30^ Prioritized genes were defined by the genome-wide TWAS and colocalization analyses described above and were not restricted to any single pre-selected locus.

### Statistical Analysis

Descriptive statistics are presented as median (interquartile range), or frequency (percentage), unless otherwise specified. Group comparisons were performed using unpaired two-sided t tests for normally distributed continuous variables, or Mann-Whitney U tests for non-normally distributed variables. Cross-sectional associations between digital biomarkers and prevalent AS were evaluated using multivariable logistic regression models, with prevalent AS as the dependent variable. Bivariate associations between continuous variables were evaluated using Pearson’s r or Spearman’s rho coefficients, as appropriate. All models were adjusted for age and sex unless otherwise specified. Effect estimates are reported as odds ratios with 95% confidence intervals. Associations between digital biomarkers and longitudinal outcomes were assessed using Cox proportional hazards regression models. Time-to-event was defined from the date of cardiovascular magnetic resonance imaging to the occurrence of AVR or death. Models were adjusted for age and sex, and results are reported as hazard ratios with 95% confidence intervals. Model discrimination was evaluated using Harrell’s concordance (C) index, with confidence intervals estimated using bootstrap resampling with r=200 repetitions. All hypothesis tests were two-sided, and a significance threshold of α of 0.05 was used unless otherwise specified. Correction for multiple testing was applied where appropriate, including false discovery rate control in transcriptome-wide association analyses. Genome and transcriptome-wide significance thresholds were applied for genetic analyses as described in the corresponding sections. Key analyses were performed using Python (v3.11).

## Data availability

The underlying data were accessed through an approved application to the UK Biobank (#71033). Following an application to the UK Biobank committee, our team was granted a waiver to export imaging and ECG data into a secure Yale university-managed server to enable inference using our deep learning-enabled algorithms. The summary GWAS statistics for our quantitative metrics, namely peak aortic velocity, DASSi and AI-ECG score of AS will be made available on Zenodo upon publication.

## Code availability

Our analytical code will be released through a link to our project’s GitHub page at the time of publication.

## Acknowledgements

This research has been conducted using the UK Biobank Resource under application number #71033.

## Author Contributions

E.K.O. conceived and supervised the study. W.L., R.B.C., D.Y., L.S.D., and P.M.C. performed the analyses and interpreted the data. W.L. and E.K.O. drafted the manuscript. All authors critically revised the manuscript for important intellectual content and approved the final version.

