## Supplementary material for "Digital phenotyping of aortic stenosis-related remodeling reveals complementary structural, electrical, and hemodynamic signatures": Online Supplement

*by*

Wenjing Luo, PhD, Ryan B. Choi, BA, Doris Yang, AB AM, Lovedeep S. Dhingra, MD, MHS, Philip M. Croon, MD, Rohan Khera, MD, MS, Evangelos K. Oikonomou, MD, DPhil

**Supplementary Figures**

**Figure S1 |** Cohort selection for cross-sectional digital biomarker analysis.
**Figure S2 |** Derivation of peak and 99$^{th}$ percentile aortic velocities from phase-contrast CMR.
**Figure S3 |** Phenome-wide association studies (PheWAS) of electrical, structural and flow digital biomarkers of the AS phenotype.
**Figure S4 |** Correlations between multimodal digital AS biomarkers and cardiac structural, functional, and electrophysiological traits.
**Figure S5 |** Prognostic value of multimodal digital biomarkers stratified by peak aortic velocity.
**Figure S6 |** Quantile-quantile (QQ) plots of the three AS-related digital remodeling phenotype GWAS.
**Figure S7 |** Convergence of digital-phenotype GWAS loci with published aortic stenosis genetics.
**Figure S8 |** Systematic colocalization for peak aortic velocity loci.
**Figure S9 |** Systematic colocalization for AI-ECG loci.
**Figure S10 |** Systematic colocalization for DASSi loci.

**Supplementary Tables (provided in** excel format in a separate spreadsheet as one table per tab)

**Table S1 |** Genome-wide significant and suggestive lead loci for each digital phenotype.
**Table S2 |** Overlap of lead loci with published aortic stenosis GWAS, with enrichment testing.
**Table S3 |** Direct look-up of lead loci in published aortic stenosis summary statistics.
**Table S4 |** Five-trait genetic-correlation matrix with two aortic stenosis references and sample-overlap intercept.
**Table S5 |** Significant transcriptome-wide association (TWAS) genes by phenotype and tissue.
**Table S6 |** Cross-study concordance of prioritized genes with a published TWAS.
**Table S7 |** Systematic colocalization across all prioritized GWAS–TWAS loci.
**Table S8 |** Valve transcriptomic contextualization of prioritized peak-velocity genes.

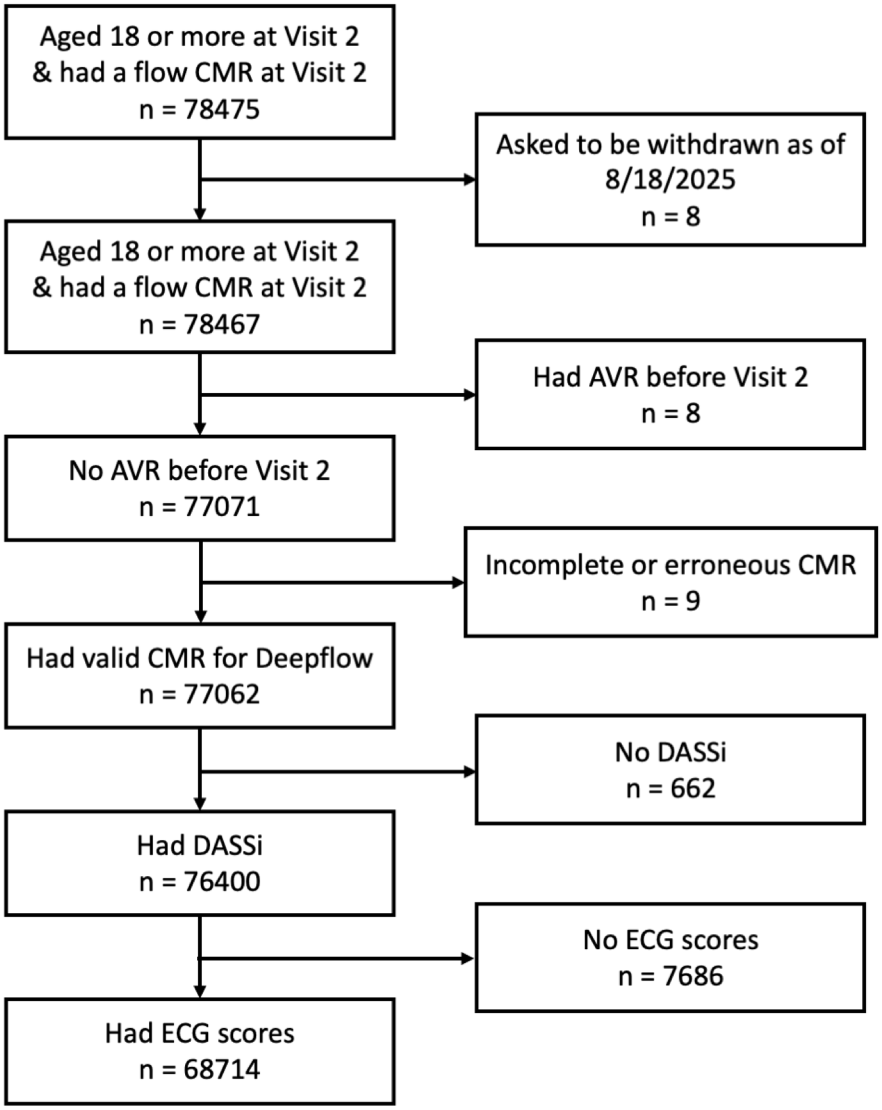


**Figure S1 | Cohort selection for cross-sectional digital biomarker analysis.** Flow diagram detailing the stepwise inclusion of UK Biobank participants with cardiovascular magnetic resonance (CMR) flow imaging at Visit 2. After exclusions for consent withdrawal, prior aortic valve replacement (AVR), incomplete CMR, or missing data, a total of 68,714 individuals had valid estimates for all three digital biomarkers: AI-enabled electrocardiogram (ECG) score, Digital Aortic Stenosis Severity index (DASSi), and peak aortic valve velocity. *Abbreviations:* AS, aortic stenosis; AVR, aortic valve replacement; CMR, cardiovascular magnetic resonance; DASSi, Digital Aortic Stenosis Severity Index; ECG, electrocardiogram.

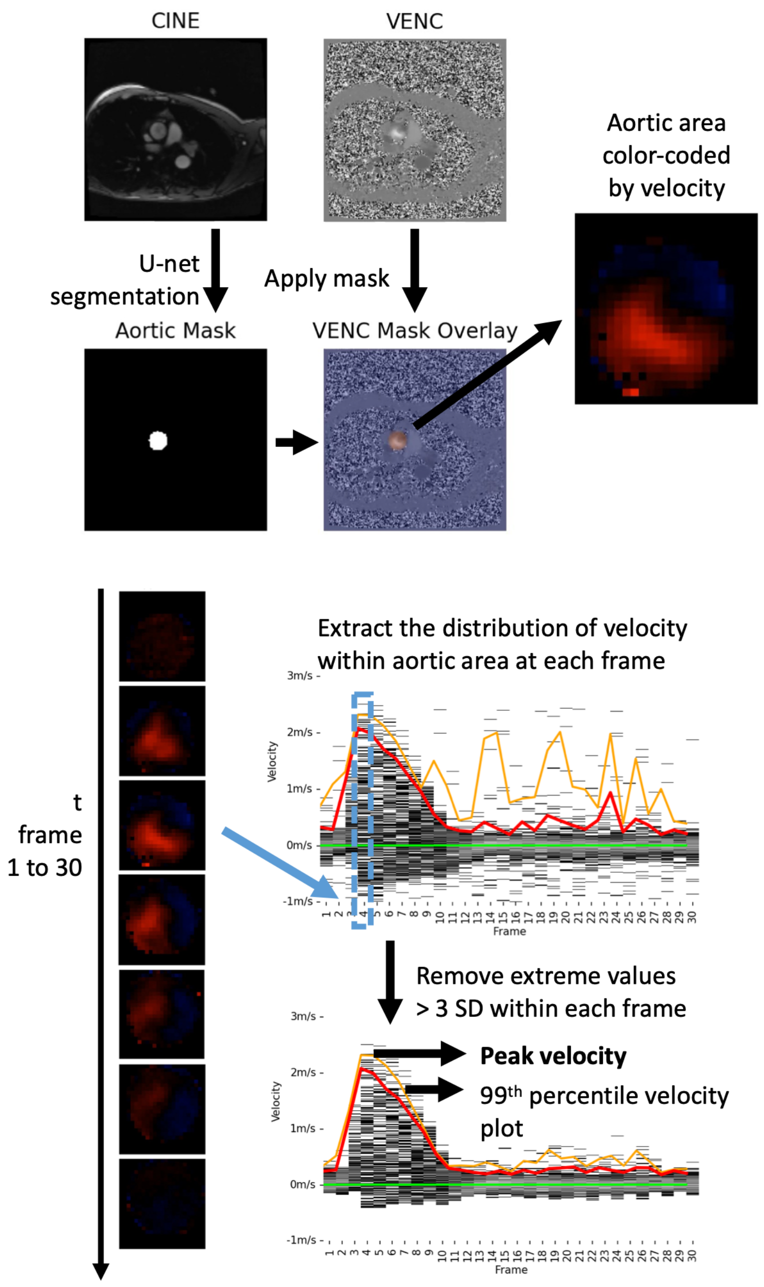


**Figure S2 | Derivation of peak and 99th percentile aortic velocities from phase-contrast CMR.** For each cardiac frame, a previously validated U-Net segmentation model was applied to cine magnitude images to delineate the aortic cross-section at the level of the sinotubular junction. The resulting aortic mask was then applied to the corresponding velocity-encoded (VENC) image to extract pixel-level velocity values within the aortic area, which were visualized using a velocity colormap (red indicating forward flow and blue indicating backward flow). Velocity distributions were obtained for each of the 30 cardiac frames and combined across the cardiac cycle. To reduce noise, velocity values exceeding three standard deviations from the frame-specific mean were excluded. The 99th percentile velocity was computed for each frame to generate the 99th percentile velocity curve, and the maximum value of this curve was defined as the peak aortic velocity. *Abbreviations:* CMR, cardiovascular magnetic resonance; VENC, velocity-encoded.

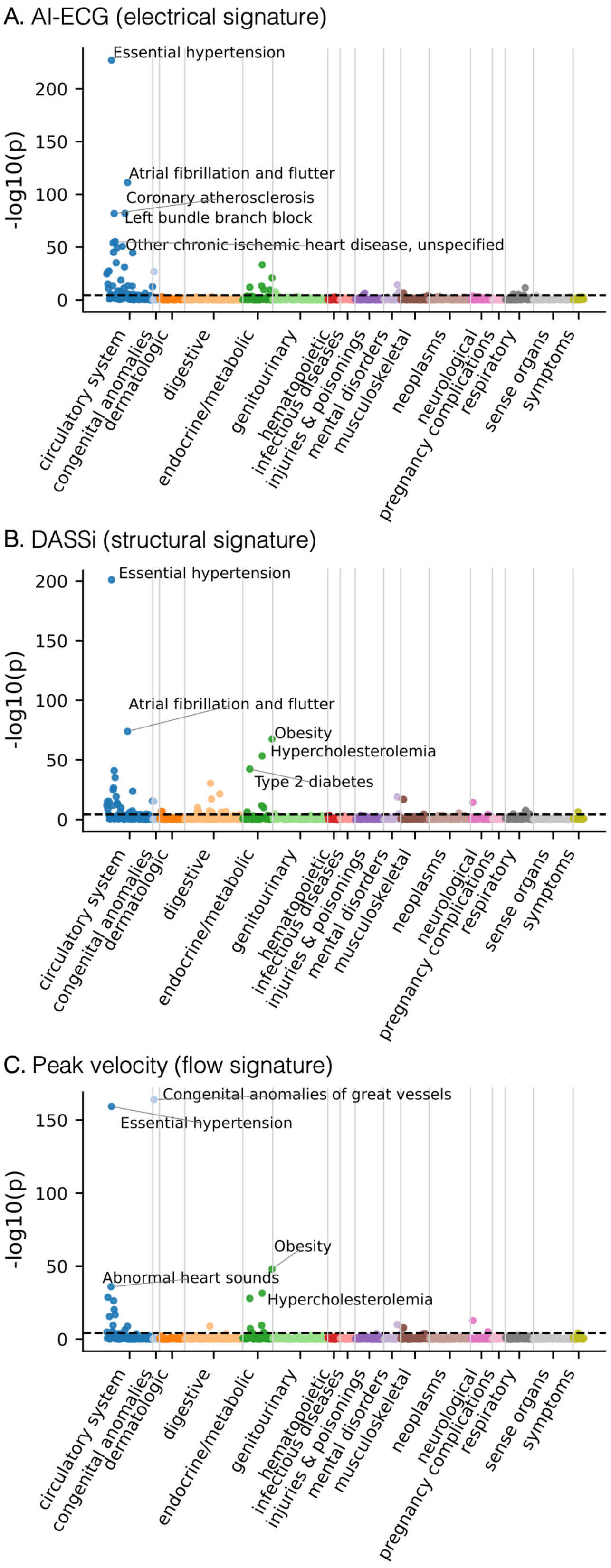


**Figure S3 | Phenome-wide association studies (PheWAS) of electrical, structural and flow digital biomarkers of the AS phenotype.** PheWAS evaluating clinical correlates of three complementary digital biomarkers of aortic stenosis (AS): **(A)** Electrophysiological signature derived from an AI-enabled ECG (AI-ECG). **(B)** Structural signature quantified by the Digital AS Severity Index (DASSi). **(C)** Flow-based signature represented by peak aortic velocity from velocity-encoded cardiovascular magnetic resonance. Panels display −log10-transformed P-values across phenotypic categories. Across modalities, digital AS remodeling-related biomarkers demonstrate predominantly cardiovascular-specific associations, with partially distinct phenotypic profiles reflecting complementary remodeling domains. *Abbreviations:* AI-ECG, artificial intelligence-enabled electrocardiogram score; AS, aortic stenosis; CMR, cardiovascular magnetic resonance; DASSi, Digital Aortic Stenosis Severity Index; PheWAS, phenome-wide association study.

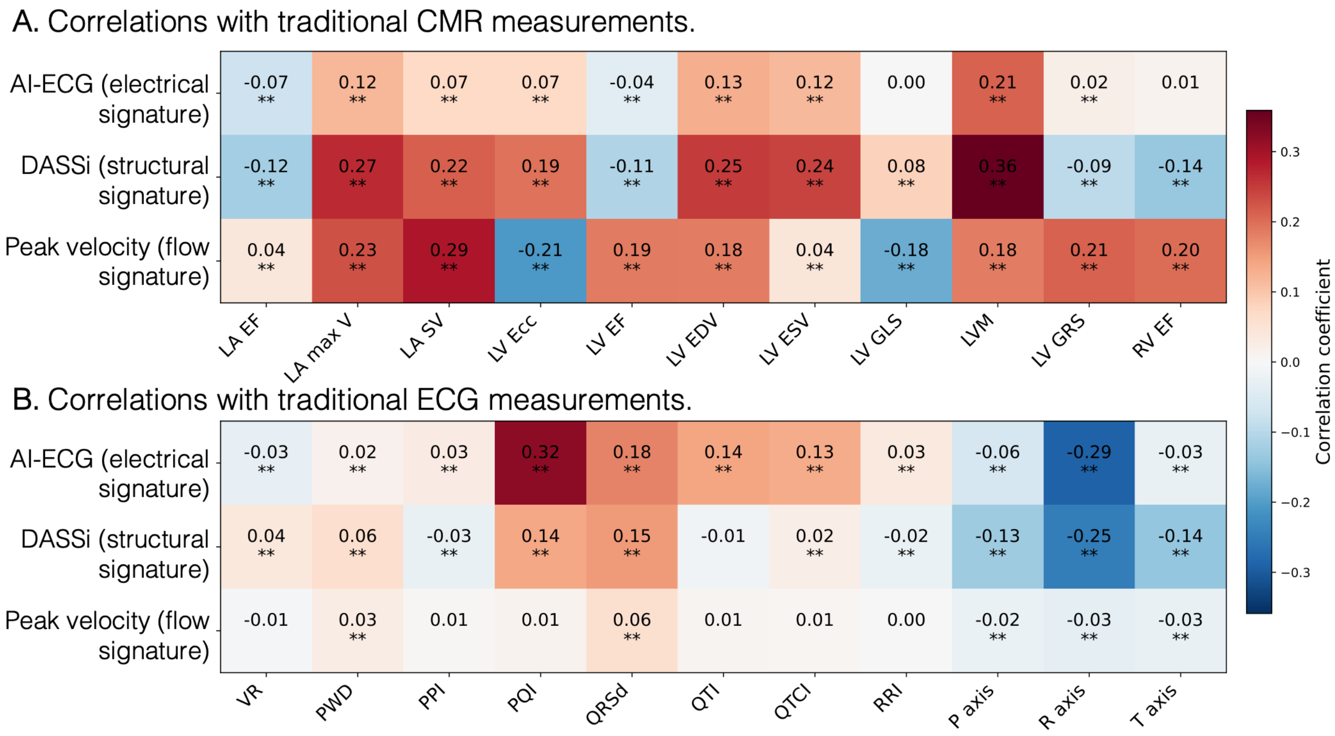


**Figure S4 | Correlations between multimodal digital AS biomarkers and cardiac structural, functional, and electrophysiological traits.** Heatmaps display pairwise Pearson correlation coefficients between three complementary digital aortic stenosis (AS) biomarkers, namely AI-enabled ECG score (ECG Score), Digital AS Severity Index (DASSi), and peak aortic velocity, and a panel of cardiac magnetic resonance (CMR)-derived (top) and electrocardiographic (ECG)-derived (bottom) quantitative traits. CMR metrics include left atrial (LA) and left ventricular (LV) structural and functional parameters (e.g., volumes, ejection fraction, strain, and mass), as well as right ventricular (RV) ejection fraction. ECG metrics include standard interval-, axis-, and variability-based measures of electrical conduction and repolarization. Color intensity reflects the magnitude and direction of correlation (red, positive; blue, negative). Asterisks denote associations significant after multiple-comparison correction ($P < 0.05$). Sample sizes for each trait are as follows: Abbreviations and sample sizes: LA EF, left atrial ejection fraction (n=32,141); LA max V, left atrial maximum volume (n=32,141); LA SV, left atrial stroke volume (n=32,141); LV Ecc, left ventricular circumferential strain (n=32,535); LV EF, left ventricular ejection fraction (n=32,584); LV EDV, left ventricular end-diastolic volume (n=32,584); LV ESV, left ventricular end-systolic volume (n=32,584); LV GLS, left ventricular global longitudinal strain (n=31,630); LVM, left ventricular mass (n=32,584); LV GRS, left ventricular global radial strain (n=32,535); RV EF, right ventricular ejection fraction (n=32,584); VR, ventricular rate (n=68,714); PWD, P-wave duration (n=65,090); PPI, P-P interval (n=40,479); PQI, PR (PQ) interval (n=38,493); QRSd, QRS duration (n=68,714); QTI, QT interval (n=40,479); QTCI, heart rate-corrected QT interval (n=40,479); RRI, R-R interval (n=40,479); P axis, P-wave axis (n=38,573); R axis, QRS axis (n=40,479); T axis, T-wave axis (n=40,479).

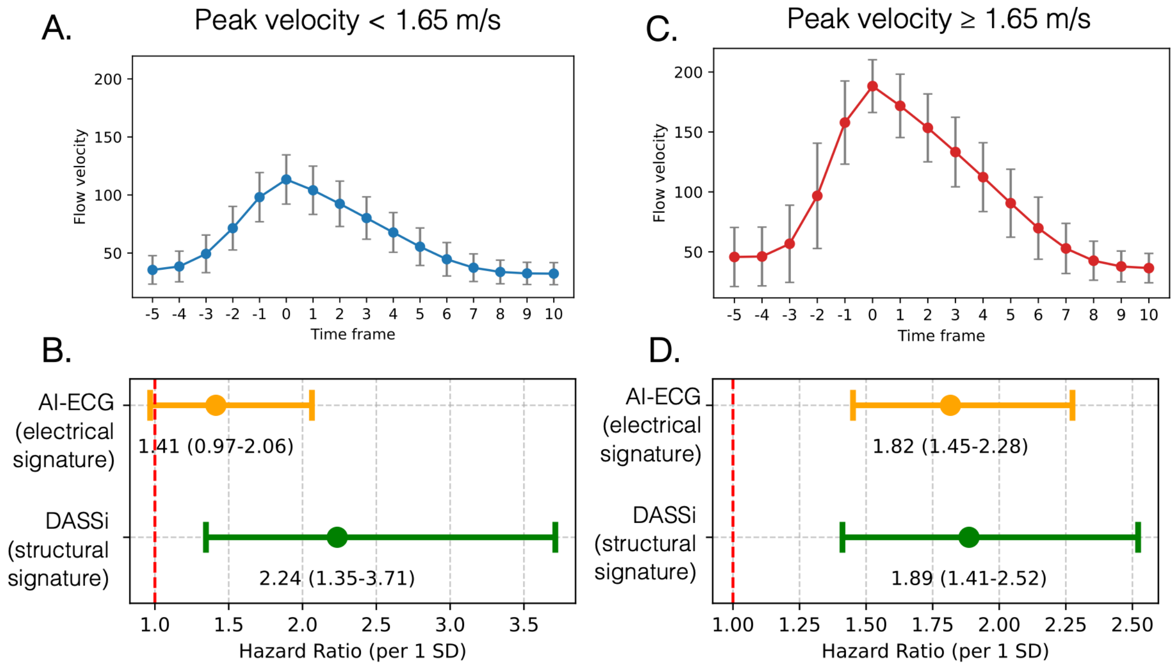


**Figure S5 | Prognostic value of multimodal digital biomarkers stratified by peak aortic velocity.** (**a**, **c**) Representative velocity-time profiles from velocity-encoded CMR in individuals with peak aortic velocity < 1.65 m/s (**a**) and ≥ 1.65 m/s (**c**); curves are the mean across individuals with error bars denoting one standard deviation, with cardiac frames temporally aligned to the frame of peak velocity (time 0). (**b**, **d**) Hazard ratios and 95% confidence intervals for incident aortic valve replacement (AVR) per one standard deviation increase in AI-ECG (electrophysiological) and DASSi (structural) biomarkers, from Cox proportional hazards models adjusted for age and sex, among individuals with lower (< 1.65 m/s; b) and higher (≥ 1.65 m/s; d) peak aortic velocity. The 1.65 m/s threshold is a population-derived CMR cut-point rather than a Doppler echocardiographic definition of mild AS, and these subgroup analyses are exploratory; AVR event counts are small, particularly in the lower-velocity subgroup (16 events). *Abbreviations:* AI-ECG, artificial intelligence-enabled electrocardiogram score; AS, aortic stenosis; AVR, aortic valve replacement; CMR, cardiovascular magnetic resonance; DASSi, Digital Aortic Stenosis Severity Index.

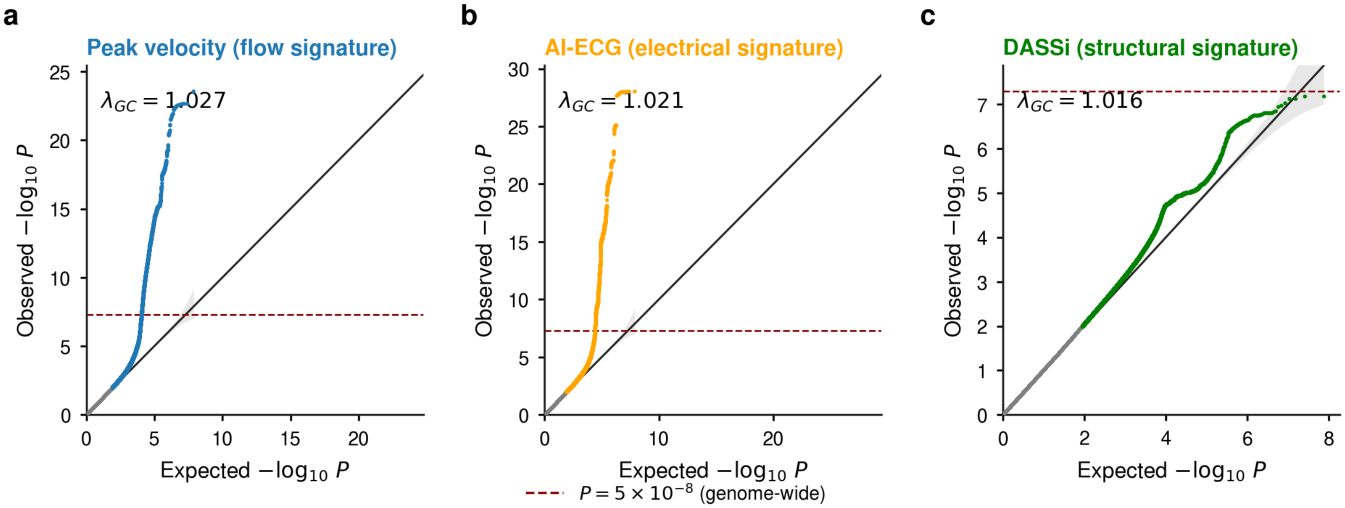


**Figure S6 | Quantile-quantile (QQ) plots for the three aortic stenosis (AS)-related digital remodeling phenotype genome-wide association studies.** QQ plots of observed versus expected −log10 P for (**a**) peak aortic velocity, (**b**) AI-ECG AS score, and (**c**) DASSi. The grey band denotes the 95% null concentration band; the genomic-control inflation factor ($\lambda_{GC}$) is shown in each panel ($\lambda_{GC}$ = 1.027, 1.021, and 1.016 for peak aortic velocity, AI-ECG, and DASSi, respectively). The dashed red line marks conventional genome-wide significance ($P = 5 \times 10^{-8}$). For all three phenotypes, departure from the null is confined to the extreme tail, consistent with polygenic signal rather than residual confounding. The corresponding Manhattan plots are shown in Fig. 4. Abbreviations: AI-ECG, artificial intelligence-enabled electrocardiogram score; AS, aortic stenosis; DASSi, Digital Aortic Stenosis Severity Index; QQ, quantile–quantile; $\lambda_{GC}$, genomic-control inflation factor.

**Peak aortic velocity**

| | Small | Kany |
|---|---|---|
| 17q21.31 | ● | – |
| *KCNRG* | – | – |
| 8p23.1 | ● | ● |
| *CDK8* | ● | – |
| *PDE3A* | ● | ● |
| *HMGA2* | ● | ● |
| *MN1* | – | ● |
| *LPA* | ● | ● |
| *OTUD7B* | ● | ● |
| *ZEB2* | ● | ● |
| *WNT4* | – | ● |
| *CHRM3* | – | – |
| *ZDHHC7* | – | – |

**AI-ECG**

| | Small | Kany |
|---|---|---|
| *VGLL2* | – | – |
| 8p23.1 | ● | ● |
| *TBX3* | – | – |
| *CAMK2D* | – | – |
| *SCN10A* | – | – |
| *SIPA1L1* | ● | – |
| *CASQ2* | – | – |
| *KCND3* | – | – |
| *BACH1* | ● | – |
| *DPT* | – | – |
| *CNOT1* | – | – |
| *NFIA* | – | – |
| *CCDC141* | – | – |

**Figure S7 | Convergence of digital-phenotype GWAS loci with published AS genetics.** Family-wise-significant lead loci ($P < 1.67 \times 10^{-8}$) for peak aortic velocity and AI-ECG, ordered by significance, and their overlap with two large published AS genetic studies (Small et al. and Kany et al.). A filled circle indicates that the locus shares an exact variant with, lies within 500 kb of, or shares a gene with a lead locus of that study; a dash indicates no overlap. Eight of the 13 peak aortic velocity loci overlap each study, compared with three (Small et al.) and one (Kany et al.) of the 13 AI-ECG loci. DASSi had no family-wise-significant locus. Inversion regions (e.g., 8p23.1 and 17q21.31) are shown as single loci. *Abbreviations:* AI-ECG, artificial intelligence-enabled electrocardiogram score; AS, aortic stenosis; DASSi, Digital Aortic Stenosis Severity Index; GWAS, genome-wide association study.

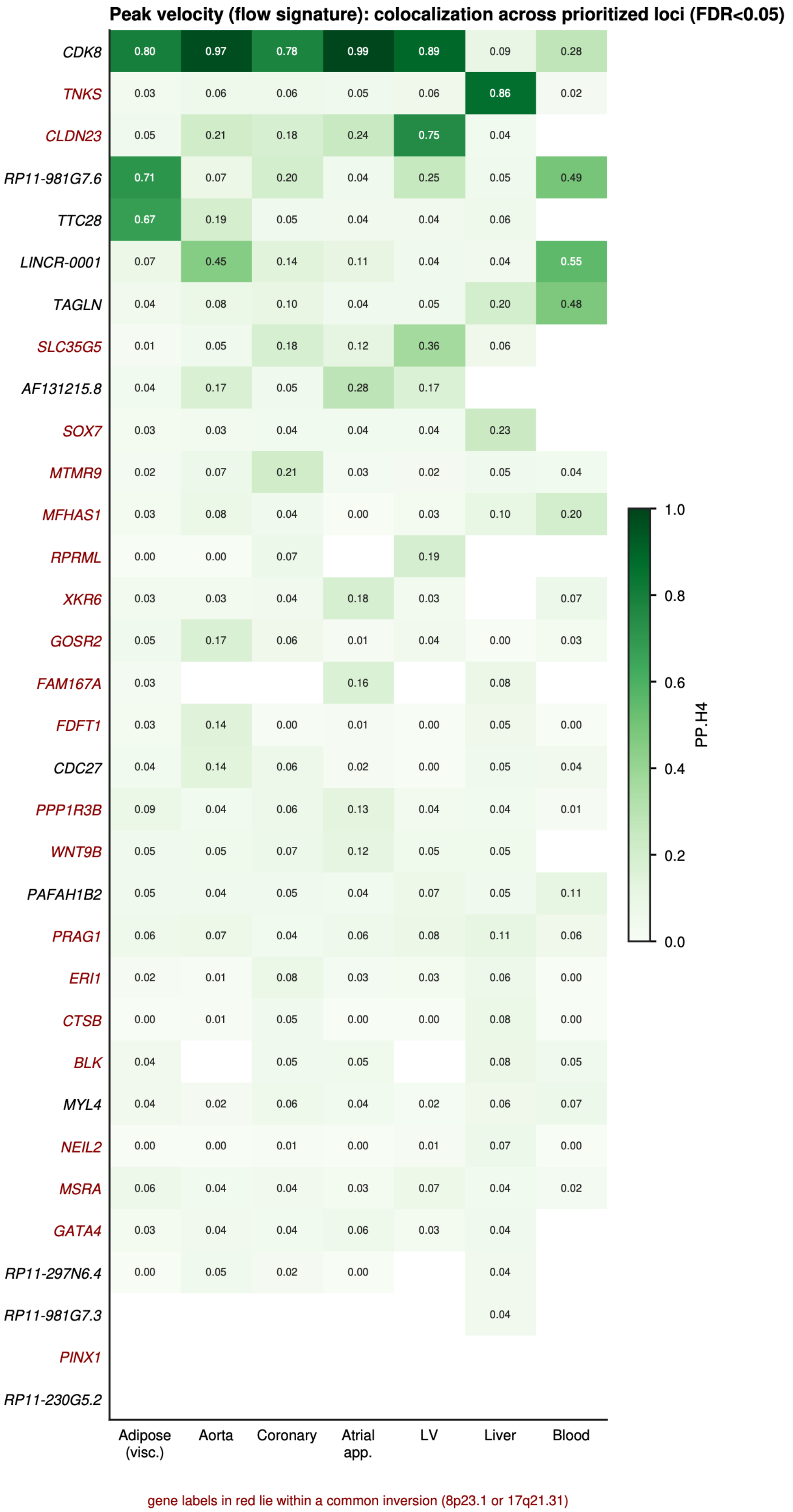


**Figure S8 | Systematic colocalization for peak aortic velocity loci.** Posterior probability of a shared causal variant (PP.H4; coloc.abf) for all prioritized GWAS × TWAS peak aortic velocity loci (TWAS false-discovery rate [FDR] < 0.05) across seven cardiometabolically relevant tissues. Rows, genes (sorted by maximum PP.H4); columns, tissues; colour, PP.H4. Both positive and negative results are shown; blank cells indicate loci with insufficient shared variants or no available cis-eQTL for that gene–tissue pair. Gene labels in red lie within common inversion polymorphisms (e.g., 8p23.1 or 17q21.31); colocalization there cannot prioritize one gene over the others the region contains, and these genes are treated as provisional. *Abbreviations:* eQTL, expression quantitative trait locus; FDR, false-discovery rate; GWAS, genome-wide association study; PP.H4, posterior probability of a shared causal variant; TWAS, transcriptome-wide association study.

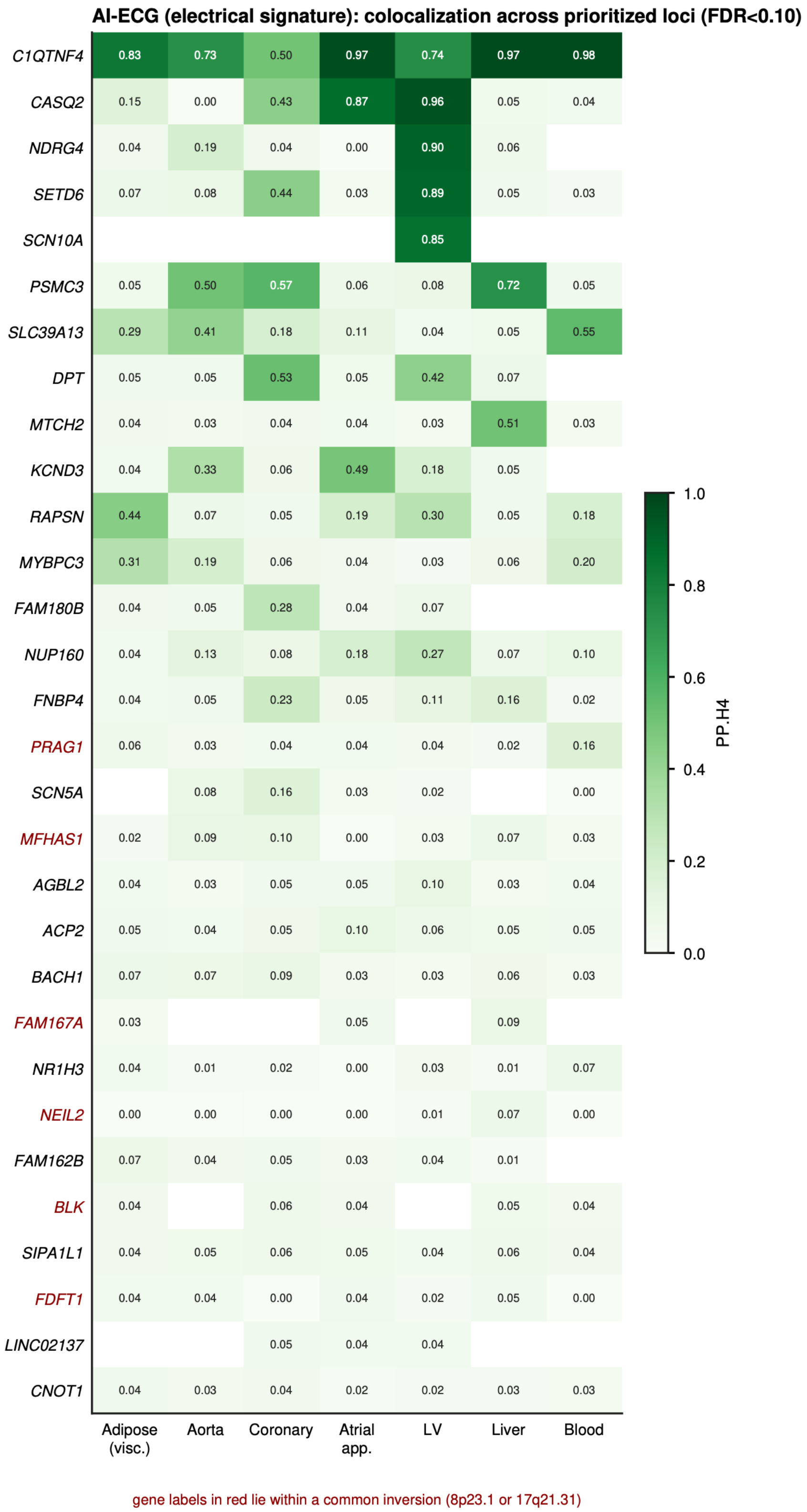


**Figure S9 | Systematic colocalization for AI-ECG loci.** As in Figure S8, for all prioritized AI-ECG GWAS × TWAS loci (TWAS FDR < 0.10) across seven tissues. *Abbreviations:* AI-ECG, artificial intelligence-enabled electrocardiogram score; FDR, false-discovery rate; GWAS, genome-wide association study; TWAS, transcriptome-wide association study.

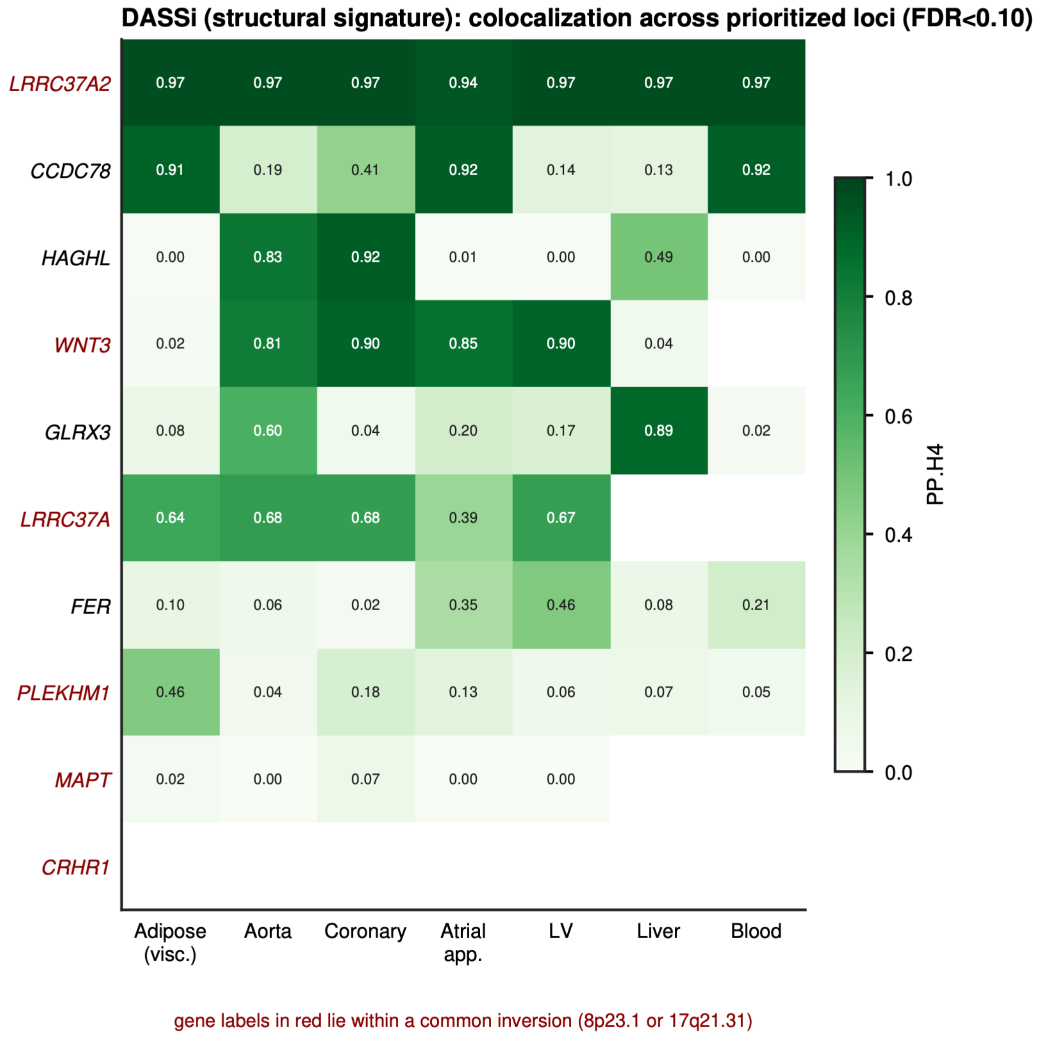


**Figure S10 | Systematic colocalization for DASSi loci.** As in Figure S8, for all prioritized DASSi GWAS × TWAS loci (TWAS FDR < 0.10) across seven tissues. Half of the colocalizing genes lie in the chromosome 17q21.31 inversion region (gene labels in red), including *LRRC37A* and *LRRC37A2*. *LRRC37A2* shows near-identical posterior probabilities across all seven tissues (PP.H4 0.94-0.97), a pan-tissue pattern consistent with inversion haplotypes, copy-number variation, or imputation effects rather than tissue-specific causal regulation, and is treated as provisional. *WNT3* is an exception, colocalizing preferentially in cardiovascular tissues (aorta, coronary artery, atrial appendage, and left ventricle) but not in adipose tissue or liver, a more tissue-specific pattern that marks it as a more plausible candidate. *Abbreviations:* DASSi, Digital Aortic Stenosis Severity Index; FDR, false-discovery rate; GWAS, genome-wide association study; PP.H4, posterior probability of a shared causal variant; TWAS, transcriptome-wide association study.
